# Cortical and subcortical structural alterations in obsessive-compulsive disorder: relationships between morphology and clinical profiles in the Global OCD study

**DOI:** 10.1101/2025.07.23.25332068

**Authors:** Niels T. de Joode, Chris Vriend, Petra J.W. Pouwels, Feng Liu, Maria C.G. Otaduy, Bruno Pastorello, Frances C. Robertson, Jonathan Ipser, Seonjoo Lee, Anton J.L.M. van Balkom, Dianne M. Hezel, Marcelo C. Batistuzzo, Marcelo Q. Hoexter, Karthik Sheshachala, Janardhanan C. Narayanaswamy, Ganesan Venkatasubramanian, Christine Lochner, Euripedes C. Miguel, Janardhan Reddy YC, Roseli G. Shavitt, Dan J. Stein, Melanie M. Wall, H. Blair Simpson, Odile A. van den Heuvel

## Abstract

**Background:** The Global OCD consortium investigated cortical and subcortical alterations in obsessive-compulsive disorder (OCD) using T1-weighted MRI, focusing on the relationship between brain morphology and clinical phenotypes, illness duration, comorbidity and medication history.

**Methods:** This preregistered study included 266 medication-free adults with OCD and 254 healthy controls. We analyzed cortical thickness, surface area, and (sub)cortical and cerebellar volumes using FreeSurfer ROI metrics and voxel-based morphometry (VBM). All analyses included age, sex, IQ, and site as covariates.

**Results:** We found no significant differences in cortical thickness, surface area, or subcortical volumes between OCD and HC groups. OCD severity correlated negatively with surface area in the left lateral orbitofrontal cortex (OFC) (p_(FDR)_=0.008), right medial OFC (p_(FDR)_=0.009), and right insula (p_(FDR)_=0.045), findings that were supported by VBM analyses. Analyses related to obsessive-compulsive (OC) symptom dimensionality showed a negative correlation between harm and aggression scores and surface area in the right medial OFC (p_(FDR)_=0.030), while sexual and religious scores positively correlated with hippocampal volume (p_(FDR)_=0.017). Compared to those without, OCD cases with comorbid depressive disorders showed a smaller surface area of the inferior parietal cortex (p_(FDR)_=0.048) and bilateral middle temporal cortices (R:p_(FDR)_=0.048; L:p_(FDR)_=0.049). No associations were found for illness duration, age of onset, or past medication use.

**Conclusions:** This study identified structural changes associated with illness severity, OC symptom dimensionality, and comorbidities. Our results emphasize the role of the OFC and insula in OCD and suggest comorbidity-specific alterations. The absence of case-control differences may stem from the inclusion of medication-free individuals and clinical heterogeneity within the OCD group.

## 1. Introduction

Previous T1-weighted structural MRI (sMRI) studies in obsessive-compulsive disorder (OCD) described alterations in cortical and subcortical morphometry. Specifically, regional alterations were observed in the cortico-striato-thalamo-cortical (CSTC) circuit as well as fronto-limbic and fronto-parietal regions(1–3). Relative to controls, adults with OCD show decreased cortical thickness in the parietal and temporal cortices(2) and lower gray matter volume (GMV) in the orbitofrontal, dorsolateral prefrontal, medial prefrontal, and inferior frontal cortices(4,5).

Subcortical volumetric alterations, notably in the thalamus and the pallidum, have also been observed. The thalamus serves a central role within the CSTC circuits. Although volumetric thalamic alterations are most pronounced in pediatric OCD (with larger volume in unmedicated children with OCD), adults with OCD show smaller thalamic volume compared to controls(1,6,7). Additionally, smaller hippocampus and amygdala, larger pallidum and altered cerebellar volumes are seen in adult OCD as compared to controls(1,8–11).

Most of the observed morphological alterations are dependent on clinical characteristics such as medication status, illness duration or severity, comorbid disorders, and obsessive-compulsive (OC) symptom dimensions(1,2,4–6,12). Medicated adults with OCD showed more pronounced and widespread thinning of the cortex as well as more evident subcortical volumetric alterations(2,6). Adult OCD cases with early onset and/or longer illness duration showed larger pallidum and smaller thalamic and amygdala volume(1,6,8,10).

Although previous mega- and meta-analyses, such as those by the ENIGMA-OCD consortium, used large pooled datasets of individual-participant data with harmonized data processing, such harmonization was limited by the use of retrospective image acquisition and clinical assessments. Thus, the relationship between structural brain changes and various clinical characteristics and medication use could not be satisfactorily examined, despite indications that these characteristics may play an important role(2).

To address these limitations, the Global OCD study was launched(13). This global initiative benefits from harmonized prospective methods for clinical phenotyping(14), neurocognitive testing(15), and state-of-the-art neuroimaging(16) in a large, ethno-culturally diverse sample of medication-free individuals with OCD and HC across five international sites(13). To identify consistent and generalizable brain markers of OCD across diverse international cohorts, we integrated multiple sMRI techniques, including FreeSurfer and voxel-based morphometry (VBM), to assess the cortical thickness, surface area, and (sub)cortical and cerebellar volumes.

The main objectives of this study are to 1) improve the current understanding of cortical and subcortical alterations in OCD compared to controls, 2) assess subcortical structures that are relevant to OCD (i.e. thalamus, amygdala, and hippocampus) using sub-segmentation methods to investigate subnucleic alterations, and 3) establish the relationship between structural alterations and clinical characteristics within the OCD sample, including OC symptom (dimension) severity, age of OCD onset, duration of illness, medication history, and comorbid depressive and anxiety disorders.

We hypothesized that compared to HC, adults with OCD would show morphometric alterations in both cortical (particularly in the prefrontal cortices) and subcortical areas (primarily thalamus, hippocampus, and amygdala). Within the OCD group, we expected that OC symptom severity, age of OCD onset, illness duration, presence of comorbid depressive and anxiety disorders, and medication history would correlate with these structural alterations.

## 2. Materials and methods

### 2.1. Participants

The Global OCD Study recruited 281 medication-free adults with OCD (18-50 years old), and 265 age-, sex-, and education-matched HCs across five research sites in São Paulo, Brazil; Bangalore, India; Amsterdam, the Netherlands; Cape Town, South Africa; and New York, U.S.A.(13). OCD was determined as the primary diagnosis using the Structured Clinical Interview for DSM-5 (SCID)(17), with a minimum Yale-Brown Obsessive-Compulsive Scale (Y-BOCS)(18,19) score of 16.

Exclusions for OCD participants included: psychotropic medication use (other than prn sleep medicines) or exposure and response prevention therapy in the prior six weeks; history of a psychotic/bipolar/anorexia nervosa/Tourette disorders; or current substance use disorder, bulimia, or chronic tic disorder. Exclusions for HCs included: a lifetime psychiatric disorder other than major depressive disorder or an anxiety disorder (if not in past year); history of psychotropic medications (other than prn sleep medications); or a first-degree relative with OCD/tic disorder. Exclusions for all included: MRI contraindications; major medical or neurological disease/head trauma; pregnancy; acute suicidality; or an intelligence quotient (IQ) below 80(13).^1^ Participants provided written informed consent, and local ethical committees at each site approved the study.

### 2.2. Clinical Measures

A comprehensive list of the clinical measures administered in this study(13) and the fidelity of core clinical assessments across five sites(14) has been published previously. We used the Y-BOCS to assess overall OCD symptom severity and the dimensional Y-BOCS (DY-BOCS)(20) to evaluate the presence and severity of distinct symptom dimensions within OCD.

Additionally, depressive and anxiety subsamples were established based on current SCID diagnoses. The depressive subsample comprised participants with current major depressive disorder (MDD) and/or persistent depressive disorder, while the anxiety subsample included those with current agoraphobia, generalized anxiety disorder, panic disorder, posttraumatic stress disorder, social anxiety disorder, or specific phobia. Also, to capture the spectrum of symptom severity, the Hamilton Anxiety Rating Scale (HAM-A)(21) and Hamilton Depression Rating Scale (HAM-D)(22) were employed to measure the severity of anxiety and depressive symptoms, respectively.

Based on previous work of our group we divided OCD participants into early-onset (<18 years) and late-onset (≥18 years) groups, determined by the earliest age at which their OCD symptoms began to significantly interfere with their activities, became time-consuming (>1 hour a day), or caused substantial distress(23). Demographic and clinical characteristics such as the duration of illness, medication history (use of selective serotonin reuptake or serotonin-norepinephrine reuptake inhibitors [SSRI/SNRI]), and IQ were recorded for all study participants.

### 2.3. Image Acquisition and Quality control

High-resolution 3D T1-weighted structural images were acquired on a 3T MRI scanner at each site following the ADNI3 protocol(24). Details on the standardization of MRI protocols across sites are described in a prior publication(16). Imaging parameters for all sites are detailed in Table S1, and preprocessing as well as quality control procedures, are outlined in the supplementary materials (Section S10.2).

### 2.4. sMRI processing and analysis

#### 2.4.1. Freesurfer cortical and subcortical

T1-weighted images were processed using FreeSurfer 7.3.2(25) for intensity non-uniformity (INU) correction, skull stripping, brain surface reconstruction, and subcortical segmentation. These procedures adhered to standardized ENIGMA protocols for harmonization of analyses and quality control across multiple sites (available at http://enigma.usc.edu/protocols/imaging-protocols/).

Cortical segmentation involved the division into 68 cortical gray matter regions (34 per hemisphere) based on the Desikan-Killiany atlas(26), including two whole-hemisphere measures. Each segmented region underwent visual inspection and statistical evaluation to identify any outliers.

The subcortical segmentation focused on 18 regions of interest (nine per hemisphere), from the ASEG atlas(27). These included seven key subcortical gray matter structures (nucleus accumbens, amygdala, caudate, hippocampus, pallidum, putamen, and thalamus) that also underwent visual inspection to ensure the accuracy of the segmentation process (Section S10.2).

#### 2.4.2. Subfield segmentation

We sub-segmented the thalamus, hippocampus, and amygdala using the ‘subfields’ modules within FreeSurfer 7.3.2. For the thalamus, we grouped the 25 nuclei into five subregions per hemisphere: anterior, lateral, ventral, intralaminar/medial, and pulvinar as previously described(6).

The hippocampal subfields were grouped into the head, body, and tail, while the amygdala was segmented into the basolateral amygdala (BLA), central medial amygdala (CMA), and cortical-like nuclei(28,29).

#### 2.4.3. Cerebellum

We first segmented the cerebellum into 28 subunits using Automatic Cerebellum Anatomical Parcellation employing U-Net Locally Constrained Optimization (ACAPULCO), that applies a deep learning algorithm to divide the cerebellum into anterior, posterior, and central divisions(30). Secondly, we used the Spatially Unbiased Infra-tentorial Template (SUIT) toolbox, in conjunction with SPM12, to generate voxel-based morphometry (VBM) maps of the cerebellar gray matter(31).

#### 2.4.4. VBM

We performed voxel-wise statistical analysis of gray and white matter volume (GMV and WMV) using the ENIGMA VBM tool developed by Matthew Kempton (https://sites.google.com/view/enigmavbm), used DARTEL to create a group template and applied a smoothing kernel size of 8mm.

### 2.5. Statistical Analysis

#### 2.5.1. FreeSurfer-based analyses

Our statistical analysis was pre-registered with the Open Science Framework (https://osf.io/bvywf), ensuring transparency and reproducibility. We adopted a dual-model approach for all sMRI analyses related to cortical thickness, surface area, subcortical volumes, and subfield segmentations, implementing site correction by employing ComBat(32,33) or by incorporating site as a random intercept in our models. For FreeSurfer-based analyses, univariate mixed model analyses were conducted in R(version 4.1.3; R Core Team) with the packages lme4(v1.1) and lmerTest(v3.1). For our primary results, we present only the findings corrected by ComBat in the results section. Random intercept models are provided in the supplementary tables (S76 to S145).

For both approaches, crude and adjusted models were executed for each region of interest (ROI). The crude model accounted for site and intracranial volume (ICV, except for cortical thickness), while the adjusted model additionally accounted for age, sex, and IQ. Diagnostic group (OCD vs HC), age of OCD onset (early vs late-onset), prior exposure to SSRI/SNRI (yes/no), current comorbid depressive disorders (yes/no), current comorbid anxiety disorders (yes/no) served as independent variables. In addition, within the OCD group, we performed mixed model regression analyses to examine associations of morphological features with the continuous variables: Y-BOCS, HAM-A, HAM-D scores, and illness duration.

To account for the large proportion of zero scores on sub-dimensions of the DY-BOCS (i.e. zero-inflated), dummy variables were added to indicate whether a participant scored >0 on a given sub-dimension(34,35). Sub-dimension scores and binary regressors were added to the model (along with age, sex and IQ). For analyses on cortical thickness, surface area, subcortical volumes, and subfield segmentations, alpha was set to p<0.05 and we corrected for multiple comparisons across the different ROIs using the False Discovery Rate (FDR; p<0.05). For models involving the DY-BOCS, FDR correction was applied within each dimension (i.e., adjusting p-values for multiple comparisons across all ROIs analyzed within that dimension). For pre-specified ROIs, we also report and interpret the uncorrected results, while FDR corrected results were interpreted for secondary ROIs.

#### 2.5.2. SPM-based analyses

For VBM and SUIT analyses, group-level investigations incorporated age, sex, IQ and ICV. Additionally, we incorporated dummy-coded site variables. We performed several two-sample group level analyses based on diagnosis, age of OCD onset, prior exposure to SSRI/SNRI, current comorbid depressive disorders, and current comorbid anxiety disorders. We also conducted second-level regression analyses, examining associations with continuous variables Y-BOCS, HAM-A, HAM-D scores, and illness duration. Within the SPM framework, addressing the high proportion of zero scores in sub-dimensions (as in the FreeSurfer analyses) was not feasible. We therefore integrated only the continuous DY-BOCS dimensional scores, and covariates of no interest as regressors for the DY-BOCS regression model. To improve comparability with previous studies, resulting T-maps for VBM and SUIT analyses were thresholded at p<.001 uncorrected at the voxel level and a minimum cluster extent of 100, after which cluster-level statistics were reported at a family-wise error corrected threshold of p=0.05(10).

### 2.7. Data Availability and Reproducibility

Upon study completion, all data from this study will be submitted to the NIMH Data Archive, an NIH-funded data repository.

## 3. Results

### 3.1. Demographics and clinical characterization

The initial sample consisted of 281 adults with OCD and 265 HC. Twelve participants (n=7 OCD; n=5 HC) were excluded after enrollment due to withdrawal of consent, failure to complete any study procedure, or later meeting study exclusion criteria. This left 274 OCD and 260 HC participants, of whom 11 (n=7 OCD; n=4 HC) did not complete the MRI with usable data. Of the 523 participants with MRI data (267 OCD; 256 HC), sMRI data from three individuals (n=1 OCD; n=2 HC) were excluded because of ventricular abnormalities, resulting in a final sample of 266 OCD patients and 254 HCs (Figure 1). OCD and HC groups were similar in sex and age distribution, but showed significant differences in estimated IQ and years of education (see Table 1 for demographic and clinical information and statistics). Characteristics of the OCD subgroups (e.g. early vs late-onset) are summarized in the supplements and Table S2-S5.

**Figure 1.**
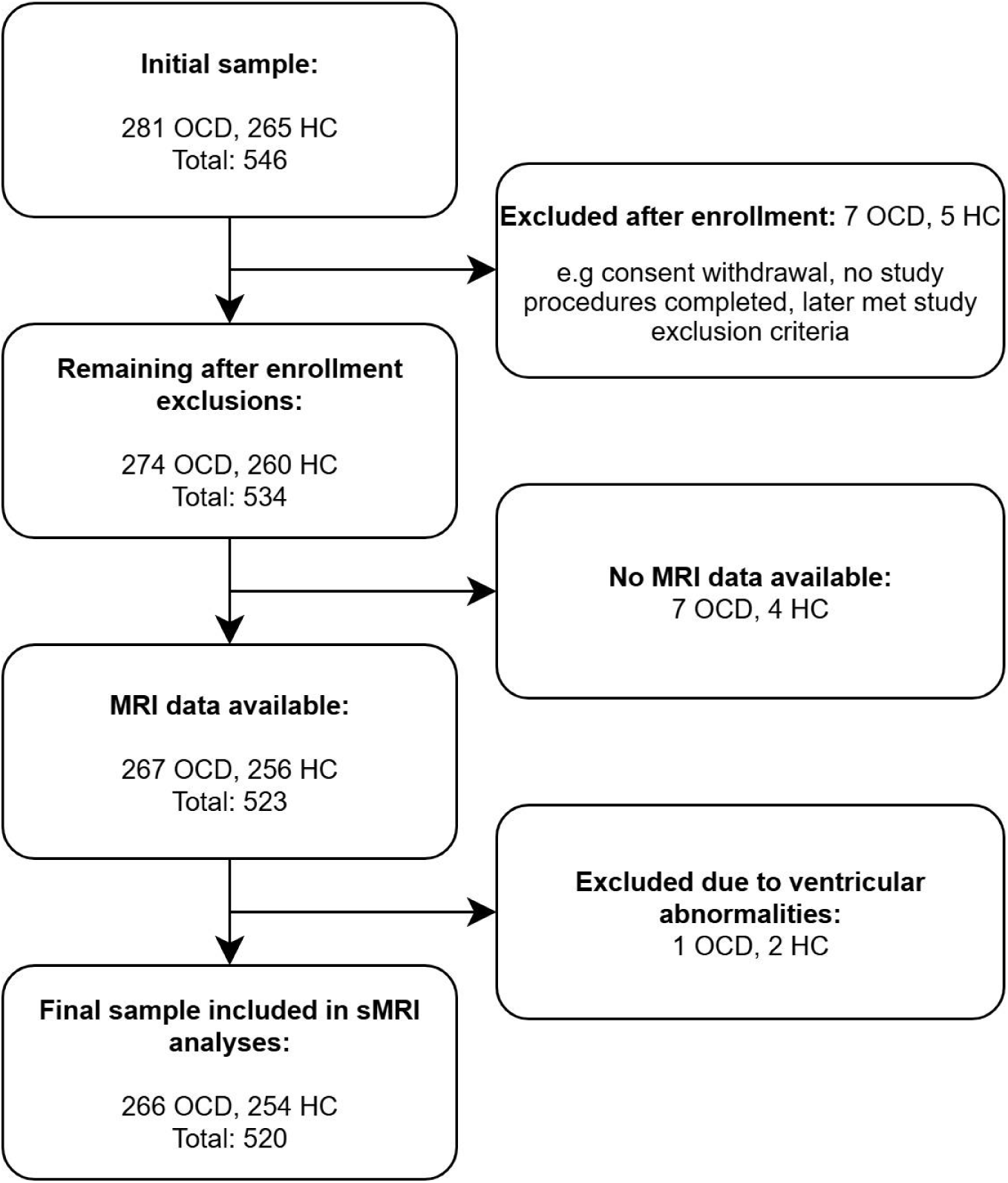
Consortium and study flowchart. Flow diagram depicting participant recruitment, inclusion, and exclusion throughout the study. The initial sample included 281 individuals with obsessive-compulsive disorder (OCD) and 265 healthy controls (HC). Twelve participants were excluded after enrollment due to consent withdrawal, failure to complete any study procedures, or meeting exclusion criteria. Of the remaining 534 participants, 11 had no MRI data available. Structural MRI (sMRI) data from an additional three participants were excluded due to ventricular abnormalities. The final sample included in the sMRI analyses consisted of 266 individuals with OCD and 254 HCs.

**Table 1.** Demographic and clinical characteristics OCD vs HC.

| Characteristic | OCD (N=266) | HC (N=254) | Statistics |
| --- | --- | --- | --- |
| **Sex (N (%))** | | | $\chi^2(1)=0.47, p=0.49$ |
| Male | 121 (42.1 %) | 107 (45.5 %) |  |
| Female | 145 (57.9 %) | 147 (54.5 %) |  |
| **Age (years)** | 29.6 (7.9) | 30 (8.2) | $t(514.88)=0.63, p=0.53$ |
| **Estimated IQ** | 104.6 (12.5) | 107.1 (12.2) | $t(517.68)=2.23, p=0.026$ |
| **Education (years)** | 15.2 (2.8) | 16 (2.5) | $t(516.28)=3.63, p<0.001$ |
| **Y-BOCS** | 24.8 (4.9) | 0.1 (0.7) | $t(275.51)=-81.33, p<0.001$ |
| **HAM-A** | 12.1 (8) | 1.5 (2.2) | $t(305.98)=-20.76, p<0.001$ |
| **HAM-D** | 8.8 (6.1) | 1 (1.8) | $t(315.02)=-20.03, p<0.001$ |
| **comorbid anxiety disorders** | 114 (42.9%) |  |  |
| **comorbid depressive disorders** | 65 (24.4%) |  |  |
| **Duration of illness<sup>a</sup>** | 12.3 (8.5) |  |  |
| **Age of OCD onset (years)** | 17.3 (7) |  |  |
| **OCD onset (N (%))<sup>a</sup>** |  |  |  |
| early-onset | 149 (56.2) |  |  |
| late-onset | 116 (43.8) |  |  |
| **Medication naïve (%)** |  |  |  |
| SSRI | 155 (58.3%) | 252 (99.2%) |  |
| SNRI | 253 (95.1%) | 254 (100%) |  |
| Benzodiazepines | 240 (90.2%) | 253 (99.6%) |  |
| Antipsychotics | 245 (92.1%) | 253 (99.6%) |  |
| Mood stabilizers | 259 (97.4%) | 254 (100%) |  |
| CBT naïve | 201 (75.6%) | 254 (100%) |  |
| **DY-BOCS** |  |  |  |
| Harm and Aggression | 5.3 (4.7) |  |  |
| Sexual and Religious | 4.4 (4.9) |  |  |
| Symmetry and Ordering | 5.8 (4.4) |  |  |
| Contamination | 6.4 (5) |  |  |
| Collecting and Hoarding | 1.3 (2.7) |  |  |

Data are presented as mean (SD) unless otherwise indicated. a=duration of illness/age of OCD onset missing for 1 patient.
Abbreviations: CBT=Cognitive Behavioral Therapy, Comorbid Anxiety=social anxiety disorder and/or generalized anxiety disorder, specific phobia, panic disorder, agoraphobia, posttraumatic stress disorder, comorbid depressive disorders=major depressive disorder or persistent depressive disorder, DY-BOCS: Dimensional Y-BOCS (severity score), HAM-A=Hamilton Anxiety Rating Scale, HAM-D=Hamilton Depression Rating Scale, SNRI=Selective Noradrenaline Reuptake Inhibitor, SSRI=Selective Serotonin Reuptake Inhibitor, Y-BOCS=Yale-Brown Obsessive-Compulsive Scale.

**Table 2.**
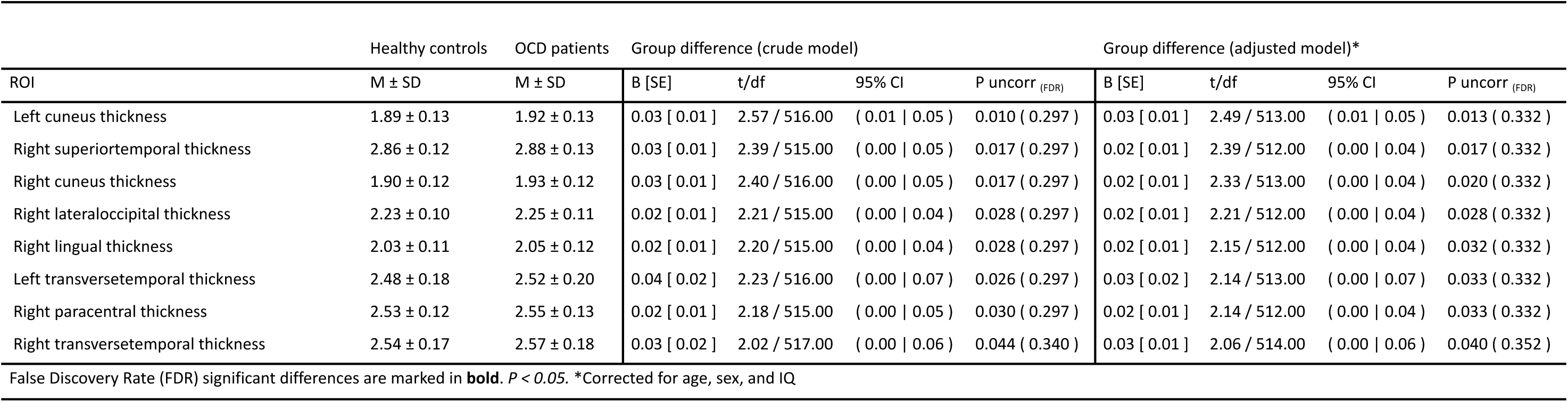
ComBat corrected results for the comparison of cortical thickness between OCD and HC.

### 3.2. OCD versus HC

Individuals with OCD versus HC demonstrated no differences in cortical thickness or surface area after FDR correction. For pre-specified ROIs, individuals with OCD demonstrated smaller surface area in the left superior frontal (B[SE]=-151.61[53.48], p=0.005, p_(FDR)_=0.324), right superior frontal (B[SE]=-126.94[50.84], p=0.013, p_(FDR)_=0.437) compared to HCs (Table S6–S7/Figure S1). Additionally, there were no between-group differences in (sub)regional subcortical or cerebellar volumes (Table S8–S12/Figure S1).

### 3.3. OCD severity

Within the OCD group, higher Y-BOCS scores were negatively associated with surface area in the left lateral orbitofrontal cortex (OFC) (B[SE]=-10.34[2.65], p_(FDR)_=0.008), right medial OFC (B[SE]=-6.88[1.86], p_(FDR)_=0.009), and right insula (B[SE]=-8.54[2.73], p_(FDR)_=0.045)(Figure 2/Table 3/Tables S13–S14/Figure S2). No associations between OCD severity and cortical thickness, subcortical volumes, or cerebellar volumes remained significant after FDR adjustment (Table S15-S19/Figure S2).

**Figure 2.**
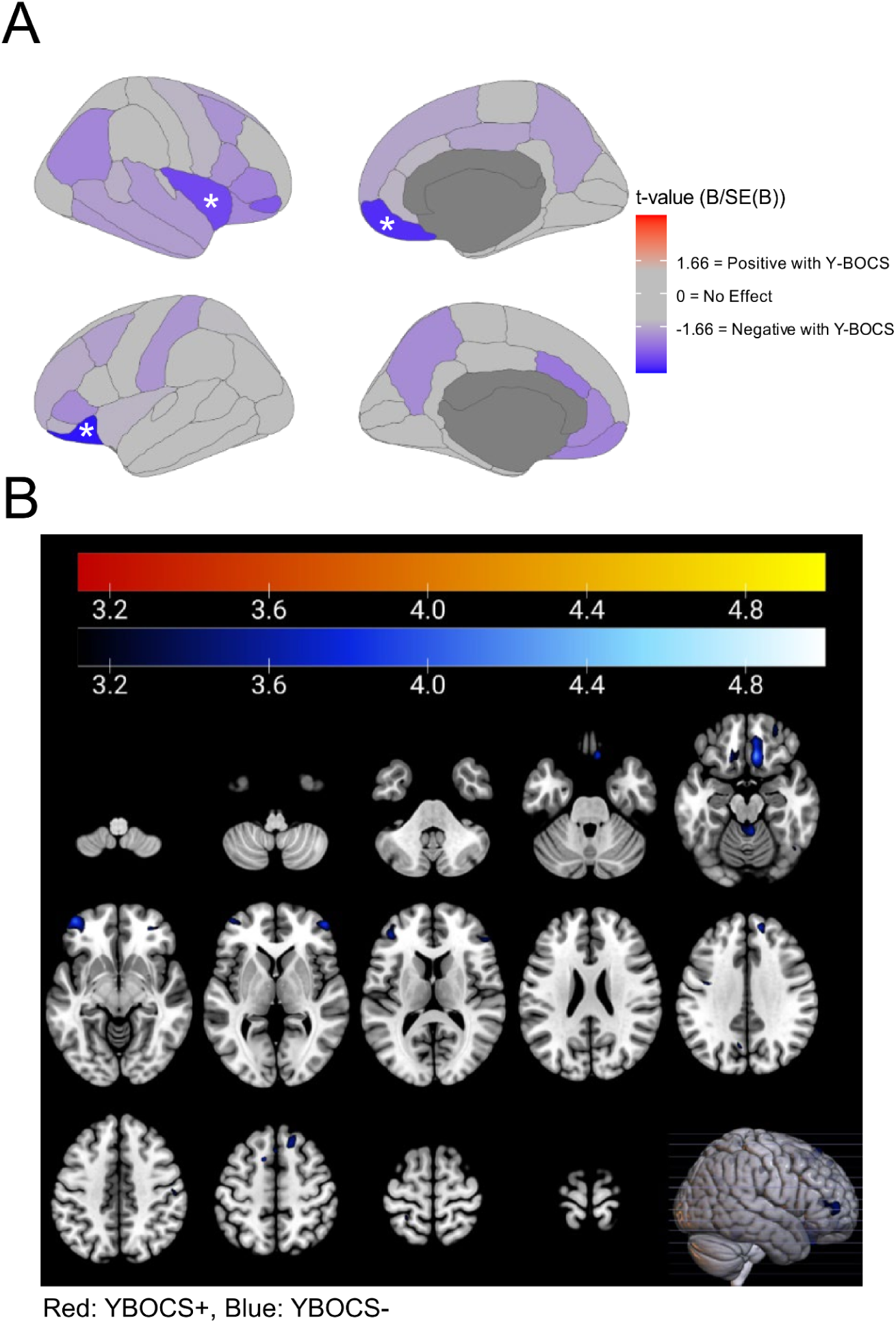
Association between morphological features and Y-BOCS Severity in OCD (N=266). A: This figure illustrates the regions of interest where surface area correlates with Y-BOCS severity in individuals with OCD. Positive associations are represented in red, with a prominent finding in the right caudal middle frontal region. Negative associations are represented in blue, and were observed for surface area and Y-BOCS score in the left lateral orbitofrontal cortex (B[SE]=-10.34[2.65], p_(FDR)_=0.008), right medial orbitofrontal cortex (B[SE]=-6.88[1.86], p_(FDR)_=0.009), and right insula (B[SE]=-8.54[2.73], p_(FDR)_=0.045). Regions in gray indicate no significant association. Color intensity reflects the strength of the standardized effect size; cooler colors denote stronger negative associations with Y-BOCS severity and significance indicated by * (p<0.05, FDR-corrected). B: Voxel-Based Morphometry (VBM) Revealing Brain Volume Decreases Associated with OCD Severity. The depicted clusters exhibit significant negative correlations with increased OCD symptom severity, with a threshold set at p <.001 uncorrected. Warm colors indicate regions where volume positively correlates with OCD severity, while cool colors show areas where volume negatively correlates with severity, depicted as t-statistic. Increased OCD severity was significantly associated with decreased GMV in a cluster spanning the right rectus, olfactory region, medial OFC and ventral striatum (t=4.32, Ke=1098, P_(FWE)_=0.022). Additional findings include decreased volume in the cerebellar Vermis 4 and 5 with higher OCD severity (t=3.99, Ke=156, P_(FWE)_=0.004).

**Table 3.** ComBat corrected results for the association between surface area and Y-BOCS scores Group difference (crude model) Group difference (adjusted model)*

| ROI | Group difference (crude model) |  |  |  | Group difference (adjusted model)* |  |  |  |
| --- | --- | --- | --- | --- | --- | --- | --- | --- |
|  | B [SE] | t/df | 95% CI | P uncorr<sub>(FDR)</sub> | B [SE] | t/df | 95% CI | P uncorr<sub>(FDR)</sub> |
| Left lateralorbitofrontal area | -10.19 [ 2.62 ] | -3.89 / 262.00 | ( -15.35 \| -5.03 ) | **<0.001 ( 0.009 )** | -10.34 [ 2.65 ] | -3.90 / 259.00 | ( -15.56 \| -5.12 ) | **<0.001 ( 0.008 )** |
| Right medialorbitofrontal area | -5.75 [ 1.86 ] | -3.09 / 263.00 | ( -9.41 \| -2.09 ) | 0.002 ( 0.075 ) | -6.88 [ 1.86 ] | -3.71 / 260.00 | ( -10.54 \| -3.22 ) | **<0.001 ( 0.009 )** |
| Right insula area | -7.68 [ 2.68 ] | -2.87 / 261.00 | ( -12.95 \| -2.41 ) | 0.004 ( 0.076 ) | -8.54 [ 2.73 ] | -3.13 / 258.00 | ( -13.92 \| -3.16 ) | **0.002 ( 0.045 )** |
| Right parsorbitalis area | -3.48 [ 1.18 ] | -2.95 / 262.00 | ( -5.81 \| -1.16 ) | 0.003 ( 0.076 ) | -3.47 [ 1.18 ] | -2.95 / 259.00 | ( -5.79 \| -1.15 ) | 0.004 ( 0.060 ) |
| Right lateralorbitofrontal area | -6.76 [ 2.79 ] | -2.42 / 263.00 | ( -12.26 \| -1.26 ) | 0.016 ( 0.208 ) | -6.54 [ 2.81 ] | -2.33 / 260.00 | ( -12.07 \| -1.01 ) | 0.021 ( 0.219 ) |
| Left caudalanteriorcingulate area | -3.74 [ 1.64 ] | -2.28 / 260.00 | ( -6.98 \| -0.51 ) | 0.024 ( 0.227 ) | -3.78 [ 1.67 ] | -2.27 / 257.00 | ( -7.07 \| -0.50 ) | 0.024 ( 0.219 ) |
| Left rostralanteriorcingulate area | -3.11 [ 1.50 ] | -2.08 / 263.00 | ( -6.06 \| -0.16 ) | 0.039 ( 0.258 ) | -3.44 [ 1.53 ] | -2.25 / 260.00 | ( -6.45 \| -0.44 ) | 0.025 ( 0.219 ) |
| Right inferiorparietal area | -18.15 [ 7.65 ] | -2.37 / 262.00 | ( -33.21 \| -3.09 ) | 0.018 ( 0.208 ) | -17.16 [ 7.70 ] | -2.23 / 259.00 | ( -32.33 \| -2.00 ) | 0.027 ( 0.219 ) |
| Left medialorbitofrontal area | -4.16 [ 1.87 ] | -2.23 / 262.00 | ( -7.83 \| -0.48 ) | 0.027 ( 0.227 ) | -4.18 [ 1.90 ] | -2.20 / 259.00 | ( -7.93 \| -0.43 ) | 0.029 ( 0.219 ) |
| Right parstriangularis area | -5.58 [ 2.73 ] | -2.05 / 263.00 | ( -10.94 \| -0.21 ) | 0.042 ( 0.258 ) | -5.68 [ 2.78 ] | -2.04 / 260.00 | ( -11.16 \| -0.21 ) | 0.042 ( 0.271 ) |
| Left precuneus area | -8.54 [ 4.14 ] | -2.06 / 262.00 | ( -16.70 \| -0.39 ) | 0.040 ( 0.258 ) | -8.60 [ 4.24 ] | -2.03 / 259.00 | ( -16.96 \| -0.24 ) | 0.044 ( 0.271 ) |
| Left parstriangularis area | -4.06 [ 2.21 ] | -1.83 / 262.00 | ( -8.42 \| 0.30 ) | 0.068 ( 0.385 ) | -4.35 [ 2.24 ] | -1.94 / 259.00 | ( -8.76 \| 0.06 ) | 0.053 ( 0.301 ) |

False Discovery Rate (FDR) significant differences are marked in **bold**. $P < 0.05$ . \*Corrected for age, sex, and IQ

**Table 4.** ComBat corrected results for the association between thalamus volumes and HAM-A Group difference (crude model) Group difference (adjusted model)*

| ROI | Group difference (crude model) |  |  |  | Group difference (adjusted model)* |  |  |  |
| --- | --- | --- | --- | --- | --- | --- | --- | --- |
|  | B [SE] | t/df | 95% CI | P uncorr<sub>(FDR)</sub> | B [SE] | t/df | 95% CI | P uncorr<sub>(FDR)</sub> |
| mean Whole thalamus ComBat | 12.47 [ 7.31 ] | 1.71 / 263.00 | ( -1.93 \| 26.87 ) | 0.089 ( 0.234 ) | 17.99 [ 7.20 ] | 2.50 / 260.00 | ( 3.82 \| 32.17 ) | **0.013 ( 0.032 )** |
| mean Ventral ComBat | 5.83 [ 3.49 ] | 1.67 / 263.00 | ( -1.03 \| 12.70 ) | 0.096 ( 0.234 ) | 8.65 [ 3.52 ] | 2.46 / 260.00 | ( 1.71 \| 15.59 ) | **0.015 ( 0.032 )** |
| mean Pulvinar ComBat | 3.31 [ 2.33 ] | 1.42 / 263.00 | ( -1.28 \| 7.90 ) | 0.156 ( 0.234 ) | 5.45 [ 2.25 ] | 2.43 / 260.00 | ( 1.03 \| 9.88 ) | **0.016 ( 0.032 )** |
| mean Anterior ComBat | 0.36 [ 0.23 ] | 1.53 / 263.00 | ( -0.10 \| 0.81 ) | 0.126 ( 0.234 ) | 0.33 [ 0.24 ] | 1.35 / 260.00 | ( -0.15 \| 0.80 ) | 0.177 ( 0.196 ) |
| mean ILM ComBat | 2.27 [ 1.90 ] | 1.19 / 263.00 | ( -1.48 \| 6.02 ) | 0.235 ( 0.235 ) | 2.46 [ 1.89 ] | 1.30 / 260.00 | ( -1.26 \| 6.18 ) | 0.193 ( 0.196 ) |
| mean Lateral ComBat | 0.41 [ 0.32 ] | 1.28 / 263.00 | ( -0.23 \| 1.05 ) | 0.203 ( 0.235 ) | 0.44 [ 0.34 ] | 1.30 / 260.00 | ( -0.23 \| 1.11 ) | 0.196 ( 0.196 ) |

False Discovery Rate (FDR) significant differences are marked in **bold**. $P < 0.05$ . \*Corrected for age, sex, and IQ

### 3.4. Obsessive-compulsive symptom dimensionality

#### 3.4.1. Cortical thickness and surface area

For cortical surface area, there was a negative association between the harm and aggression dimension and surface area in the right medial OFC (B[SE]=-13.11[3.68], p_(FDR)_=0.030). Uncorrected results are provided in the supplements (Section 11.2)(Table S20-S21).

#### 3.4.2. Subcortical

The contamination dimension score correlated negatively with the accumbens area (B[SE]=-5.77[1.96], p=0.0036, p_(FDR)_=0.042). The sexual and religious dimension score correlated positively with hippocampal head (B[SE]=10.64[3.86], p_(FDR)_=0.017), body (B[SE]=5.24[2.23], p_(FDR)_=0.033), and whole hippocampal volumes (B[SE]=16.96[6.37], p_(FDR)_=0.017)(Section 11.2)(Table 5 and S22-S26).

**Table 5.** ComBat corrected results for the association between hippocampus volumes and DY-BOCS dimensions Group difference (crude model) Group difference (adjusted model)*

| ROI | DY-BOCS dimension | Group difference (crude model) |  |  |  | Group difference (adjusted model)* |  |  |  |
| --- | --- | --- | --- | --- | --- | --- | --- | --- | --- |
|  |  | B [SE] | t/df | 95% CI | P uncorr<sub>(FDR)</sub> | B [SE] | t/df | 95% CI | P uncorr<sub>(FDR)</sub> |
| mean head ComBat | Sexual and Religious | 10.64 [ 3.86 ] | 2.75 / 246.00 | ( 3.03 \| 18.25 ) | **0.006 ( 0.017 )** | 10.95 [ 3.90 ] | 2.81 / 243.00 | ( 3.27 \| 18.63 ) | **0.005 ( 0.012 )** |
| mean whole hippocampus ComBat | Sexual and Religious | 16.96 [ 6.37 ] | 2.66 / 246.00 | ( 4.41 \| 29.51 ) | **0.008 ( 0.017 )** | 17.83 [ 6.44 ] | 2.77 / 243.00 | ( 5.14 \| 30.51 ) | **0.006 ( 0.012 )** |
| mean body ComBat | Sexual and Religious | 4.98 [ 2.20 ] | 2.27 / 246.00 | ( 0.65 \| 9.31 ) | **0.024 ( 0.032 )** | 5.04 [ 2.23 ] | 2.26 / 243.00 | ( 0.65 \| 9.42 ) | **0.024 ( 0.033 )** |

False Discovery Rate (FDR) significant differences are marked in **bold**. $P < 0.05$ . \*Corrected for age, sex, and IQ

### 3.5. OCD onset

In comparing early-onset and late-onset OCD, no differences in cortical thickness, surface area, or (sub)regional subcortical volumes were observed after FDR correction (Table S27-S33/Figure S5).

### 3.6. Comorbidity

#### 3.6.1. Cortical thickness and surface area

Individuals with OCD with current comorbid depression showed no differences in cortical thickness to those without. However, OCD cases with (versus without) depressive disorder comorbidity showed smaller surface area in several regions: the right inferior parietal cortex (B[SE]=-275.50[85.34], p_(FDR)_=0.048), right middle temporal cortex (B[SE]=-135.78[42.07], p_(FDR)_=0.048), and left middle temporal cortex (B[SE]=-129.75[41.90], p_(FDR)_=0.049). Individuals with (versus without) comorbid anxiety disorder showed no differences in cortical thickness and surface area (Table S34-37/Figure S3+S4).

#### 3.6.2. Subcortical and cerebellar volume

No differences in subcortical or cerebellar volumes remained after FDR correction when comparing OCD individuals with versus without depressive or anxiety disorder comorbidity (Table S38-47/Figure S3+S4).

### 3.7. Past medication exposure

Forty-three percent of participants had a history of SSRI/SNRI use, with an average of 3.48±4.25 years since last use. There were no significant differences in cortical thickness or surface area between participants with OCD with or without prior SSRI/SNRI use (Table S48-S49). No between-group differences were found in subcortical, subfield, or cerebellar volumes (Tables S50–S54).

### 3.8. Voxel-Based Morphometry

VBM analyses are provided in the supplementary materials (section 11.3/Table S146-152). Briefly, increased OCD severity was significantly associated with decreased GMV in a large cluster spanning the right rectus, olfactory region, medial OFC and ventral striatum (t=4.32, Ke=1098, p_(FWE)_=0.022, MNI_peak_=8,27,-21). In the SUIT analysis, a significant cluster (t=3.99, Ke=156, p_(FWE)_=0.004, MNI_peak_=2,-56,-15) demonstrated decreased cerebellar volumes with increased OCD severity in the cerebellar vermis lobules 4 and 5.

Longer duration of OCD illness was positively associated with increased GMV in the right superior and middle frontal gyri (t=5.5, Ke=1114, p_(FWE)_=0.021, MNI_peak_=18,21,50) and the right fusiform gyrus (t=4.32, Ke=920, p_(FWE)_=0.043, MNI_peak_=21,-70,-10). SUIT analysis revealed significant negative associations with cerebellar volume for longer duration of illness, including left cerebellum lobule 8 and adjacent areas (t=4.32, Ke=341, p_(FWE)_<0.001, MNI_peak_=-30,-57,-60), and the right cerebellum lobule 8 (t=4.29, Ke=1212, p_(FWE)_<0.001, MNI_peak_=28,-67,-48).

Individuals with OCD and comorbid depressive disorders exhibited significantly increased cerebellar volumes compared to those without comorbidity in left crus 1-2, and lobule 8, and 7b (t=4.56, Ke=516, p_(FWE)_<0.001, MNI_peak_=-37,-67,-37); the left cerebellum lobule 6 (t=4.36, Ke=142, p_(FWE)_=0.006, MNI_peak_=-33,-49,-23). Further significant clusters were observed in the right cerebellum lobules 4 and 5, the vermis, and left cerebellum lobule 6 (t=4.12, Ke=207, p_(FWE)_=0.001, MNI_peak_=5,-58,-9); and (t=4.11, Ke=181, p_(FWE)_=0.001, MNI_peak_=2,-70,-37) within vermis lobule 8 and left cerebellum lobule 8.

### 3.9. Exploratory analyses

Exploratory analyses of illness duration, anxiety and depressive symptom severity are provided in the Supplementary materials (Section 11.4). Briefly, higher HAM-A scores were associated with smaller right rostral anterior cingulate surface area (B[SE]=-2.73[0.76], p_(FDR)_=0.028). Greater thalamus volume, including the ventral region and pulvinar, was observed in individuals with higher HAM-A scores (B[SE]=17.99[7.20], p_(FDR)_=0.032; B[SE]=8.65[3.52], p_(FDR)_=0.032; B[SE]=5.45[2.25], p_(FDR)_=0.032)(Figure 3). No significant associations were found for illness duration or depressive severity (as measured by HAM-D scores) after FDR correction.

**Figure 3.**
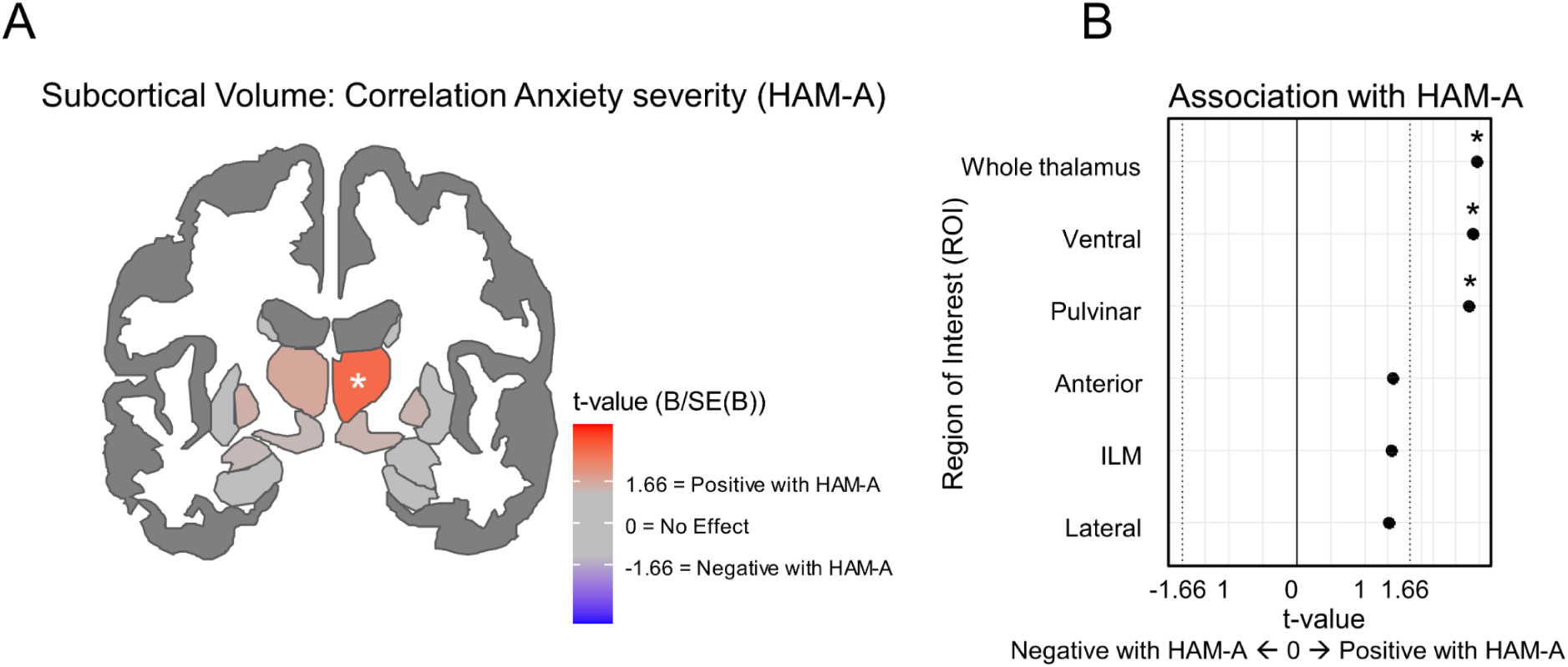
Subcortical volume associations with anxiety severity in OCD and Impact of comorbidity (N=266). A: Axial view of subcortical brain regions showing areas of significant correlation with anxiety severity (HAM-A) in OCD patients. The red area indicates a positive correlation, while the blue area indicates a negative correlation. B (Left): Subfield segmentation demonstrating the association of subcortical volumes with HAM-A scores in OCD patients. B (Right): Subfield segmentation comparing subcortical volumes in OCD patients with and without anxiety comorbidity.Scatter plot demonstrating the association of subcortical volumes with HAM-A scores in OCD patients. Black dots show the t-value of the association for each region. Significance indicated by * (p<0.05, FDR-corrected).

## 4. Discussion

This work, undertaken by the Global OCD consortium, investigated cortical and subcortical structural alterations in medication-free adults with OCD, focusing on the relationship between brain morphology and a diverse range of clinical characteristics. We did not observe case-control differences in CSTC and parietal regions, in contrast to previous studies(1,2,6,10,36,37). Our most prominent results indicated structural alterations related to illness severity, symptom dimensionality, severity of anxiety symptoms, and comorbid depressive disorders. We found more severe OCD to be related to lower surface area in the OFC and right insula, findings that were corroborated by subsequent VBM analyses. Comorbid depressive disorders were associated with smaller surface areas in the inferior parietal and middle temporal cortices, while higher anxiety levels were associated with bigger thalamic volumes. No effects of past medication use or age of OCD onset were observed.

Despite previous research, including ENIGMA-OCD and OCD Brain Imaging Consortium (OBIC) studies, indicating widespread morphometric differences in OCD compared to controls (1,2,6,10,36,37), our current study in medication-free adults with OCD did not replicate these case-control differences. Our focus on medication-free individuals might help explain the absence of case-control differences observed in our study. Indeed, previous work found that case-control differences become less pronounced(10,36), or disappear altogether in medication-free individuals(1,2). For example, Boedhoe et al. (2017/2018) showed that only medicated participants with OCD exhibited significantly thinner cortices in the frontal, temporal, parietal, and occipital regions, alongside higher pallidum volume and lower hippocampal volume. Conversely, unmedicated OCD individuals did not show these morphological differences compared to controls(1,2). Similarly, a mega-analysis by de Wit et al. (2014) found that certain gray matter alterations, such as lower middle frontal gray matter volume, were more pronounced in medicated cases compared to unmedicated ones(10).

Our most striking finding was the negative association between OCD severity and surface area in orbitofrontal regions. Although the OFC is involved in OCD, its association with severity is not consistently reported by larger-scale studies(38,39). It does however align with specific observations made by Venkatasubramanian and colleagues(40). In their study, significant negative correlations were noted with Y-BOCS scores and local gyrification index (LGI) of the right lateral and left medial OFC. They also observed that Y-BOCS insight score negatively correlated with the LGI of the left medial OFC, something we were unable to do due to limited variation in insight scores in our sample. Our VBM results clearly emphasized and confirmed the negative association with surface area, and fit with the role of the OFC in OCD(10,39).

We found dimension-specific associations involving frontal (e.g., harm and aggression dimension) as well as limbic (e.g., hippocampal) morphology, consistent with previous evidence that distinct OCD symptom dimensions may map onto partially separate orbitofrontal–striatal circuits(4,35,41–43). Specifically, higher harm and aggression scores have been linked to reduced gray matter in the amygdala and temporal lobe(4,41) and to altered connectivity in frontal striatal and limbic circuits(35). Previous studies also observed higher amygdala activation and limbic connectivity in cases with predominant sexual and religious symptoms(35,42) and that higher sexual and religious symptoms scores are associated with poorer outcomes in behavioral therapy(12,44). Although dimension-based research is complex, as individuals often show symptoms in multiple symptom dimensions, our findings underscore the neurobiological rationale for pursuing symptom-specific approaches in understanding and treating OCD(12).

Our findings regarding comorbidity diverge from previous FreeSurfer-based mega-analyses(2,45). We observed a smaller surface area in the right inferior parietal cortex and middle temporal cortices in OCD cases with current depressive comorbidity versus without. Also we observed an association with HAM-A severity and thalamic volume. The anxiety-comorbid group had a higher incidence of early-onset OCD, and larger thalamic volumes are known to be associated with childhood onset(1,6,46). However, given that thalamic alterations associated with childhood-onset OCD typically normalize by adolescence/young adulthood(46), our results suggest these thalamic volume increases are related to comorbidity rather than childhood-onset effects. Our VBM analysis found no differences in cortical regions related to comorbidity, in contrast to de Wit et al. (2014). Yet, we noted that cerebellar volume was higher in OCD patients with depressive comorbidity, particularly in the left cerebellum posterior lobules and vermis, regions known for their roles in emotion and affect regulation(47). Such cerebellar alterations, potentially indicative of depressive comorbidities, are consistent with existing research showing higher cerebellar volume in MDD(48–50).

Recent work from the Global OCD study, using other MR modalities conducted in the same sample, supports and extends our findings. Resting-state functional MRI revealed associations between clinical characteristics and connectivity patterns, with sexual and religious symptoms linked to stronger frontolimbic and sensorimotor connectivity, comorbid depression to reduced thalamo-sensorimotor connectivity and cerebellar hyperconnectivity, and anxiety severity to widespread functional hyperconnectivity(REF). The same clinical characteristics were also associated with variation in gray matter morphology presented here. Diffusion-weighted imaging similarly showed no case-control differences but revealed alterations related to age of onset, including lower posterior thalamic radiation integrity and reduced network efficiency in late-onset OCD(23). Together, these findings highlight the added value of a multimodal imaging approach with large, deeply phenotyped, and harmonized samples to identify neural alterations associated with clinical profiles in OCD.

Lastly, we observed no morphological differences between individuals with and without a history of SSRI/SNRI use. Previous ENIGMA-OCD and OBIC studies noted thinner cortex, smaller hippocampus and bigger pallidum in currently medicated patients(1,2,36). This discrepancy may stem from our exclusive focus on OCD individuals who were unmedicated not only at scanning but also for at least six weeks in advance. Although 43% of our participants had a history of SSRI/SNRI use, we observed no medication effects and the time since last SSRI/SNRI use (3.48±4.25 years) did not influence the results (data not shown). Although some studies have reported subcortical volume changes after 12 weeks of SSRI treatment(51–53) the effects of discontinuation on morphology remain understudied. It is possible that SSRI-induced changes reverse after discontinuation, which may explain the non-significant medication-related findings in our study. Future research should therefore examine the short- and long-term impact of SSRI/SNRI use and discontinuation on brain structures in OCD patients to better understand these possible transient effects.

This study has several strengths. First, the diverse and large sample size and the use of deep phenotyping and harmonized imaging acquisition protocols makes the Global OCD study the largest harmonized data collection effort in the field of OCD to date(13,16). In addition to its size, a unique strength of this sample is that all participants with OCD were unmedicated at the time of scanning, with over half being medication-naive. Secondly, the combined application of both FreeSurfer and VBM methodologies allowed for assessment of multiple morphological features simultaneously. Combining these methods helped in better capturing the complexity of brain morphology in OCD and showed that results from different methods supported each other, thereby strengthening our findings(54–56). Thirdly, the study employed a dual statistical approach using both ComBat and mixed effects models to adjust for site effects. In ENIGMA, ComBat has been shown to enhance statistical significance and power over conventional mixed effect models(33). Despite not finding a marked difference between mixed effects models and ComBat, given their similar confidence intervals, the rigorous application of these statistical methods lends further credibility to the findings.

Our study had some limitations. Firstly, for some OCD subgroup comparisons we observed subtle demographic variations potentially affecting our outcomes. For example, in the early-vs. late-onset OCD comparison, individuals with early-onset OCD were significantly younger, had a longer illness duration, and exhibited higher estimated IQ scores, potentially complicating the detection of morphological distinctions related to OCD onset. Secondly, we employed an ROI-based FreeSurfer parcellation approach using the Desikan-Killiany atlas. Atlas-based parcellations average across larger areas and might miss localized alterations(57–59). We used VBM to detect focal alterations, but this method focuses more on GMV alterations than on surface based features. Future studies can use a vertex-based analysis for more precise identification of focal effects in surface-based features(56).

Taken together, this study advances our understanding of OCD, by identifying structural alterations associated with severity, OC symptom dimensionality, and comorbidities but not with past medication use or age of illness onset. Our results not only reinforce the role of the insula and OFC in OCD, but also suggest comorbidity-specific alterations in OCD.

## 5. Acknowledgements

The authors would like to thank all participants of this study and the following study team members from each site for their vital contributions to this project: Marines Joaquim, RN; Maria Alice de Mathis, PhD and Daniel L.C. Costa, MD PhD (Brazil), Ton van Balkom, MD PhD; Neeltje Batelaan, MD PhD (Netherlands), Loche Manuel, Hons; Clara Marincowitz, MSc (South Africa), Rachel Middleton B.A.; Gabrielle R. Messner B.A.; Gabriella Restifo-Bernstein, B.A.; Page Van Meter, PhD; Sarah Rose, B.A.; Yael R. Stovetsky B.A. (U.S.A). Preliminary results were presented in the form of a poster at the Society of Biological Psychiatry annual meeting in Toronto; Canada, April 24-26, 2025. A preprint of this article was deposited at medRxiv (URL) prior to submission to the journal.

## 10. Supplementary FIGURES

**Figure S1:**
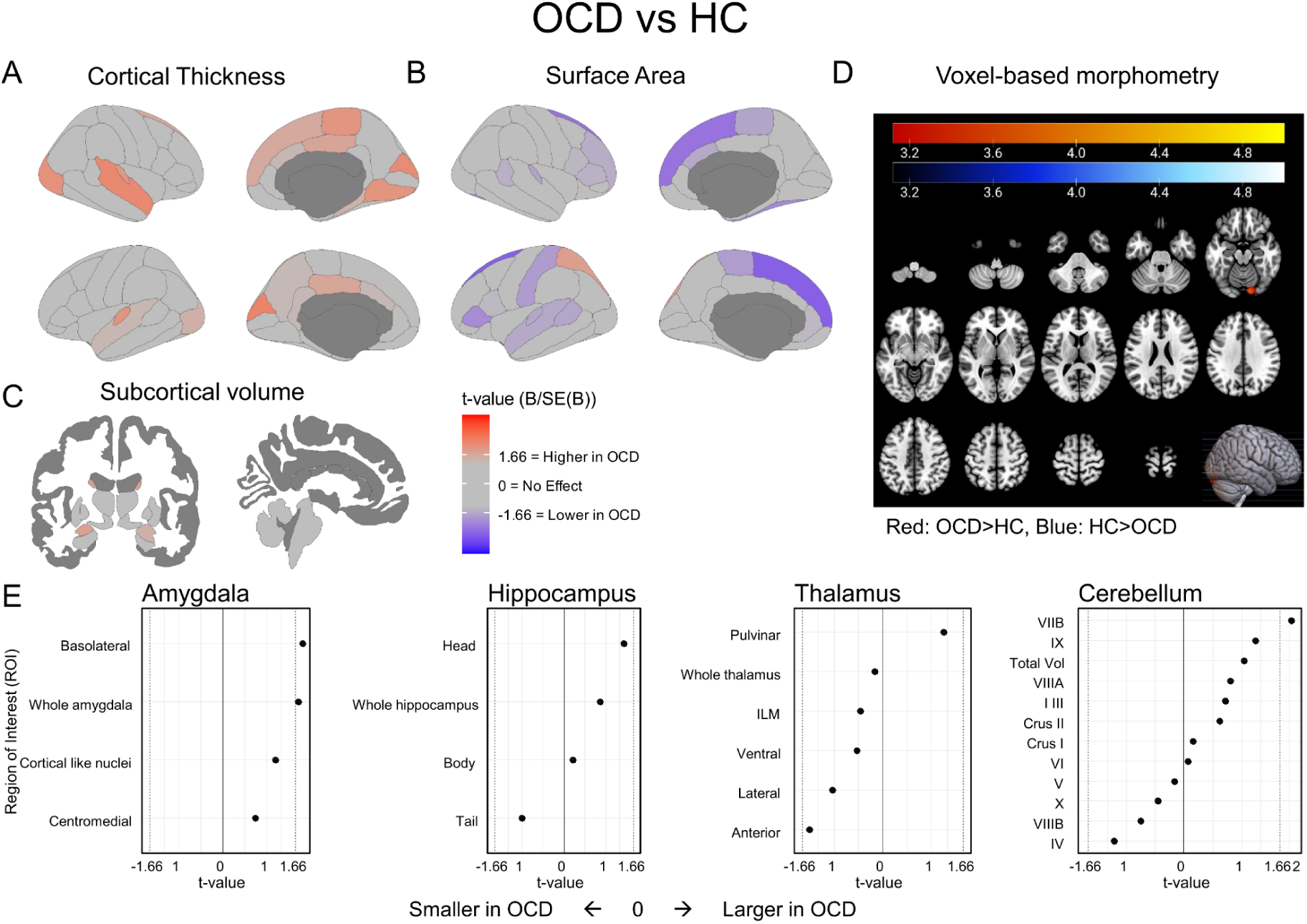
ComBat-Corrected FreeSurfer and VBM findings for comparison between individuals with OCD (N=266) and Healthy Controls (N=254). A: Cortical thickness maps with t-statistics from ComBat-corrected mixed models. B: Surface area maps with t-statistics from ComBat-corrected mixed models. C: Subcortical volume images with t-statistics from ComBat-corrected mixed models. D: Voxel-based morphometry results, shown at p<0.001 (uncorrected). E: Freesurfer subfield segmentations with t-statistics from ComBat-corrected mixed models; significance indicated by * (p<0.05, FDR-corrected).

**Figure S2:**
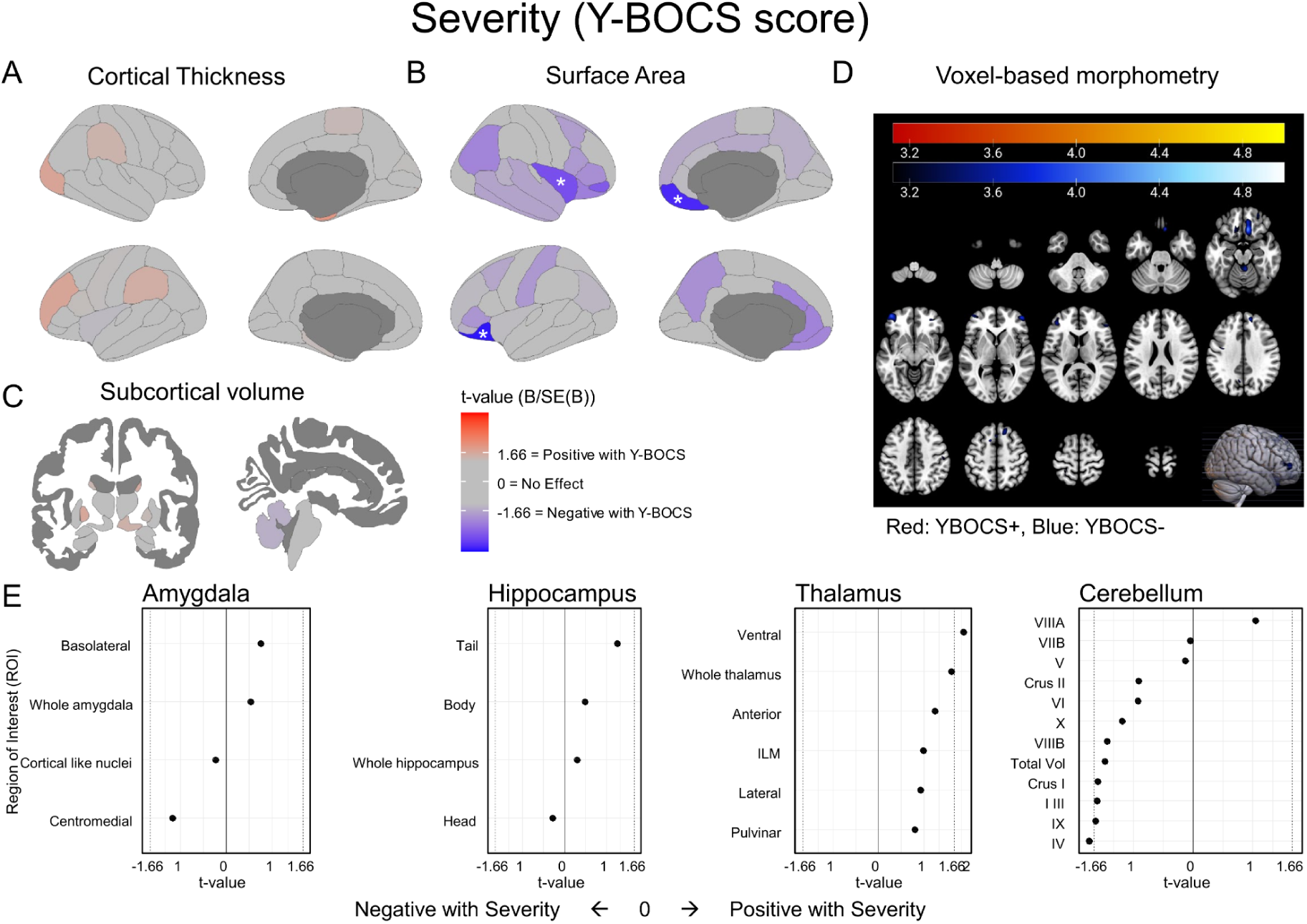
ComBat-Corrected findings for association between FreeSurfer and VBM measures and OCD severity (N=266). A: Cortical thickness maps with t-statistics from ComBat-corrected mixed models. B: Surface area maps with t-statistics from ComBat-corrected mixed models. C: Subcortical volume images with t-statistics from ComBat-corrected mixed models. D: Voxel-based morphometry results, shown at p<0.001 (uncorrected). E: Freesurfer subfield segmentations with t-statistics from ComBat-corrected mixed models; significance indicated by * (p<0.05, FDR-corrected).

**Figure S3:**
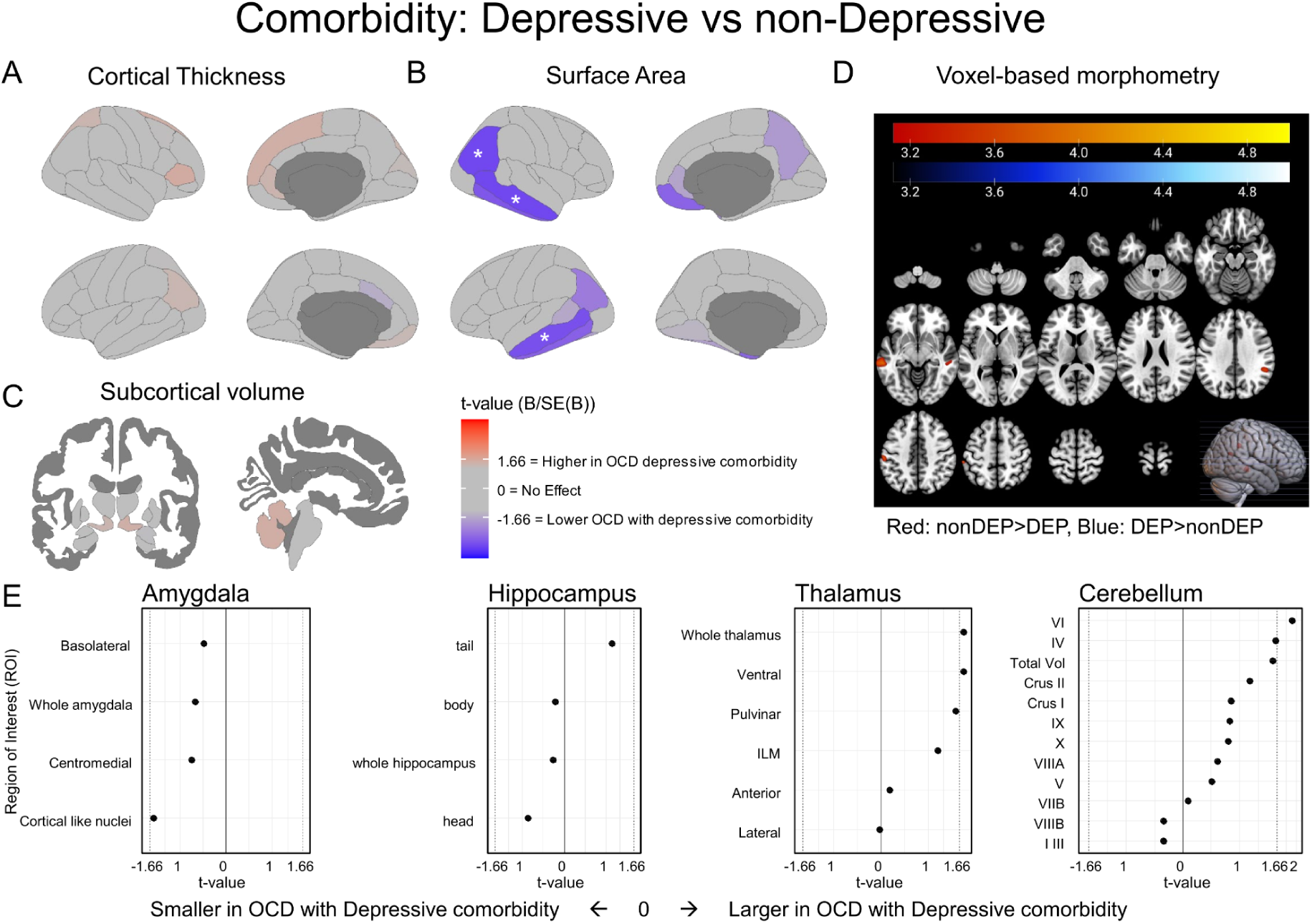
ComBat-Corrected FreeSurfer and VBM findings for comparison between OCD cases with (N=65) vs without (N=201) depressive comorbidity. A: Cortical thickness maps with t-statistics from ComBat-corrected mixed models. B: Surface area maps with t-statistics from ComBat-corrected mixed models. C: Subcortical volume images with t-statistics from ComBat-corrected mixed models. D: Voxel-based morphometry results, shown at p<0.001 (uncorrected). E: Freesurfer subfield segmentations with t-statistics from ComBat-corrected mixed models; significance indicated by * (p<0.05, FDR-corrected).

**Figure S4:**
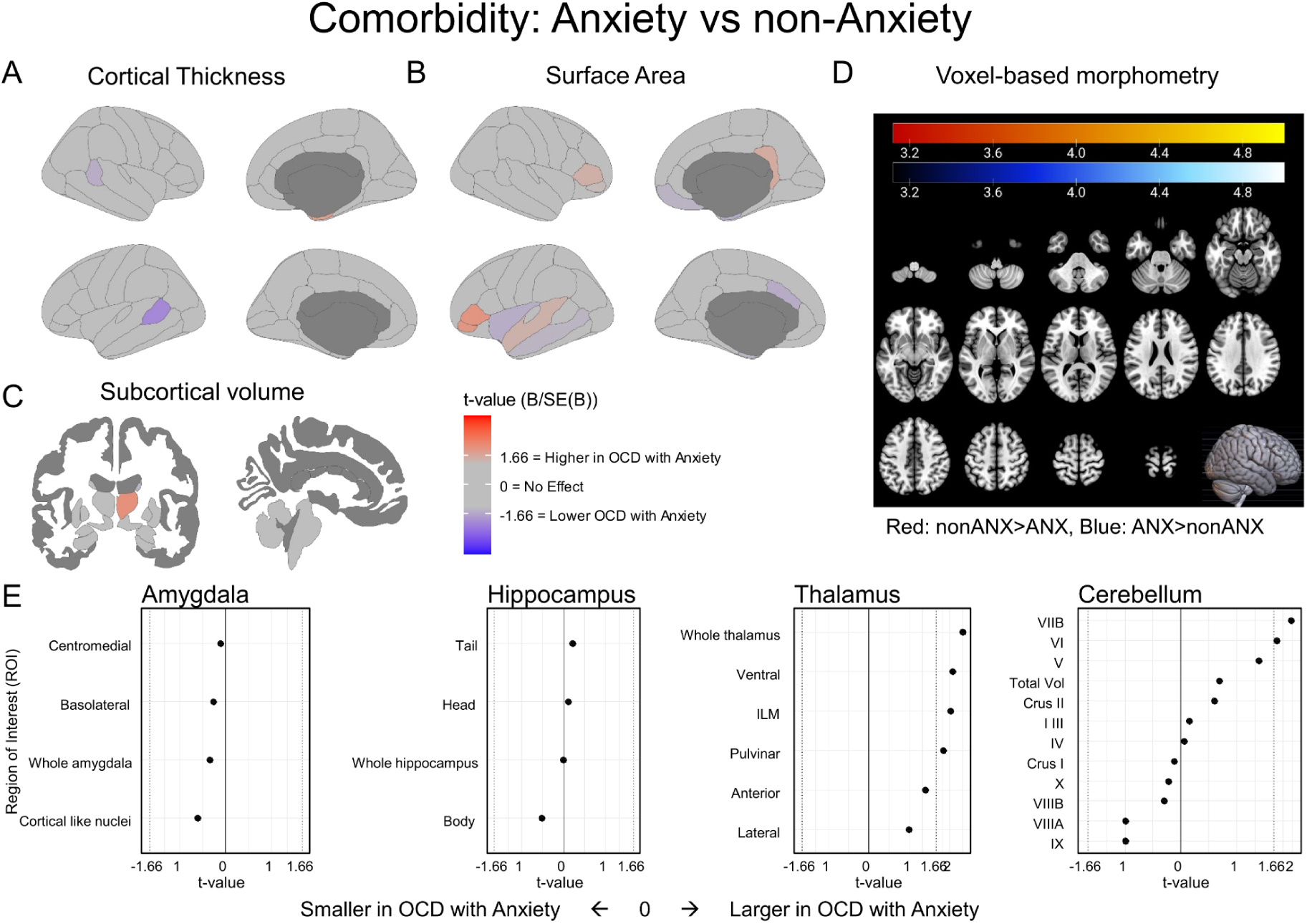
ComBat-Corrected FreeSurfer and VBM findings for comparison between OCD cases with (N=114) anxiety vs without (N=152) anxiety comorbidity. A: Cortical thickness maps with t-statistics from ComBat-corrected mixed models. B: Surface area maps with t-statistics from ComBat-corrected mixed models. C: Subcortical volume images with t-statistics from ComBat-corrected mixed models. D: Voxel-based morphometry results, shown at p<0.001 (uncorrected). E: Freesurfer subfield segmentations with t-statistics from ComBat-corrected mixed models; significance indicated by * (p<0.05, FDR-corrected).

**Figure S5:**
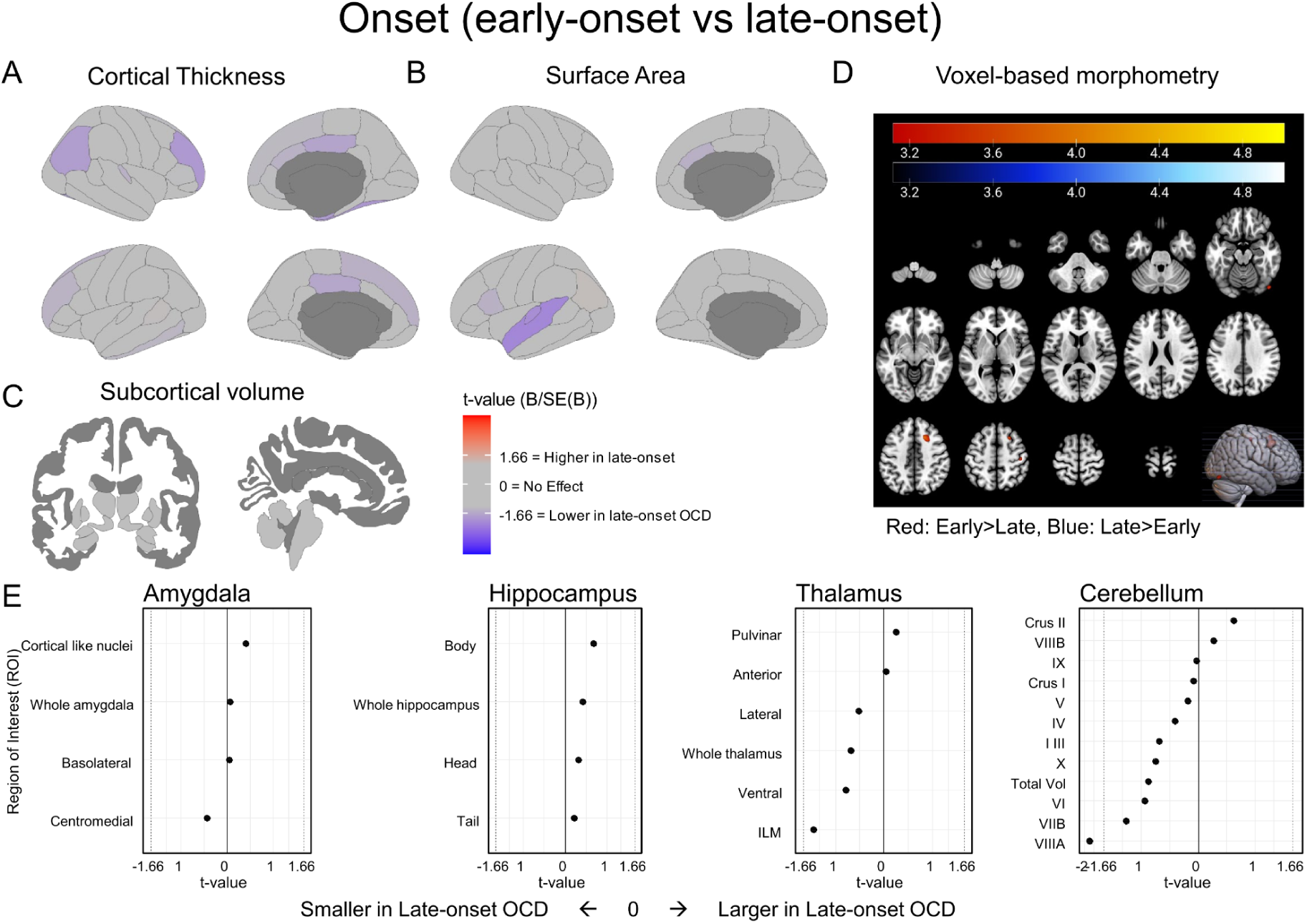
ComBat-Corrected FreeSurfer and VBM findings for comparison between OCD cases with early (N=149) vs late (N=116) onset. A: Cortical thickness maps with t-statistics from ComBat-corrected mixed models. B: Surface area maps with t-statistics from ComBat-corrected mixed models. C: Subcortical volume images with t-statistics from ComBat-corrected mixed models. D: Voxel-based morphometry results, shown at p<0.001 (uncorrected). E: Freesurfer subfield segmentations with t-statistics from ComBat-corrected mixed models; significance indicated by * (p<0.05, FDR-corrected).

**Figure S6:**
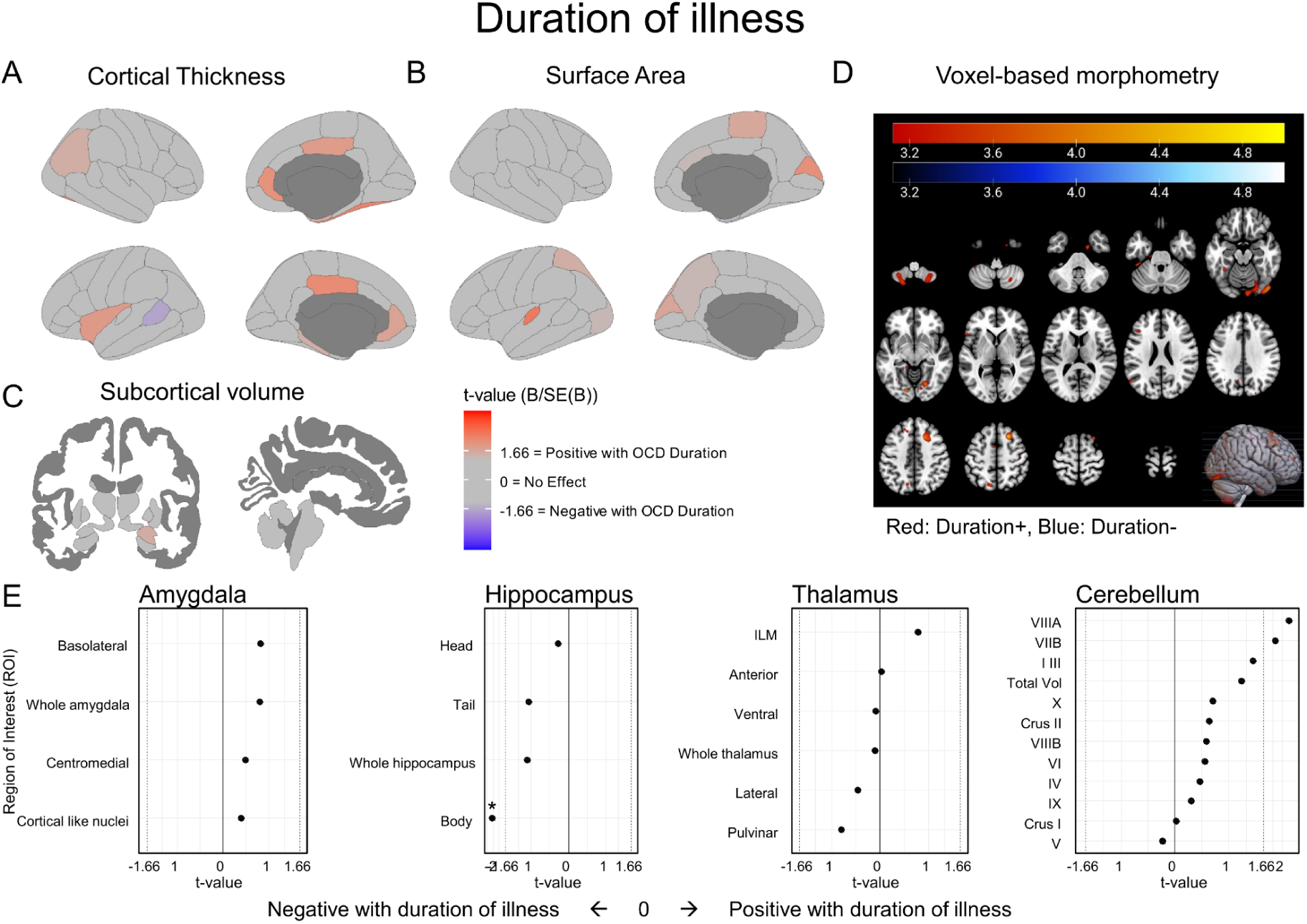
ComBat-Corrected findings for association between FreeSurfer and VBM measures and duration of illness (N=266). A: Cortical thickness maps with t-statistics from ComBat-corrected mixed models. B: Surface area maps with t-statistics from ComBat-corrected mixed models. C: Subcortical volume images with t-statistics from ComBat-corrected mixed models. D: Voxel-based morphometry results, shown at p<0.001 (uncorrected). E: Freesurfer subfield segmentations with t-statistics from ComBat-corrected mixed models; significance indicated by * (p<0.05, FDR-corrected).

## Footnotes

1 Minor exceptions were made in 13 cases (6 OCD, 7HC) with IRB notice.

