## Supplemental materials for "Cortical and subcortical structural alterations in obsessive-compulsive disorder: relationships between morphology and clinical profiles in the Global OCD study"

### 1. Supplementary METHODS

#### *1.1. Image Acquisition and Processing*

Harmonized multimodal MRI protocols were employed across all five sites using a 3.0 Tesla (T) whole-body scanner, equipped with a 32-channel phased-array head coil, except for the U.S. site which utilized a 48-channel head coil (Table S1). While scanning protocols were harmonized, minor deviations occurred due to differences in scanner vendors and models across sites. These variations and the harmonization efforts are detailed in our previous work (13,16).

#### *1.2. Quality control and visual inspection*

To ensure consistency and standardization across multiple sites, we utilized the ENIGMA consortium's standardized protocols for analyses and quality control procedures. This protocol set forth criteria for identifying outliers, defining them as cases where morphometric measurements fell > 3 standard deviations away from the group mean. We applied this criteria to cortical thickness, surface area, subcortical volumes, and subfield segmentations.

For any subject with one or more regions flagged as outliers, the corresponding segmentations underwent visual inspection using the Freeview software. This step was critical to ascertain the accuracy of the segmentation and to determine if the data of the subject should be partially or completely excluded from the analysis. It is important to note that in the FreeSurfer sub-analyses for cortical thickness, surface area, volume, and subfield segmentations, a varying number of regions failed to meet quality assurance standards and did not pass visual inspection, leading to differences in the total number of participants considered per region or morphometric analysis.

### 2. Supplementary RESULTS

#### 2.1. Demographics and clinical characterization of subsamples

Demographic and clinical information is summarized in Table 1. OCD and HC groups were similar in sex ($\chi2(1)=0.47$, $p=0.49$) and age ($t(514.88)=0.63$, $p=0.53$), but showed significant differences in estimated IQ ($t(517.68)=2.23$, $p=0.026$) and years of education ($t(516.28)=3.63$, $p<.001$).

The late-onset OCD group was on average older (mean age 31.2 vs 28.2 years, $t(248.95)=-3.12$, $p=0.002$) and had a shorter illness duration (mean duration 7.8 vs 15.8 years, t(262.33)=8.8, p<.001) compared with the early-onset group. Additionally, the late-onset group had a lower estimated IQ (mean IQ 101.8 vs 106.8, t(246.97)=3.26, p=0.0013) and a higher age of OCD onset (mean age of OCD onset 23.4 vs 12.5 years, t(176.31)=-18.68, p<.001). There was also a significant difference in comorbid anxiety (48.3% in early-onset vs 35.3% in late-onset, $\chi 2(1)$=3.98, p=0.046)(Table S2).

For SSRI/SNRI naïve (N=151) and SSRI/SNRI non-naïve (N=115) OCD participants significant differences were found in Y-BOCS scores (mean 25.6 vs 24.2, t(239.75)=-2.3, p=0.022), HAM-A scores (mean 13.4 vs 11.1, t(247.47)=-2.39, p=0.018), and HAM-D scores (mean 9.8 vs 8, t(227.05)=-2.41, p=0.017), with the SSRI/SNRI non-naïve group showing higher scores. Additionally, comorbid depressive conditions were more prevalent in the SSRI/SNRI non-naïve group (31.3% vs 19.2%, $\chi 2(1)$=4.54, p=0.033)(Table S3).

In the comparison of OCD patients with and without depressive comorbidity, significant differences were observed in the distribution of sex, with a higher percentage of females in the OCD with depressive comorbidity group (72.3%) compared to the OCD without depressive comorbidity group (48.8%, $\chi 2(1)$=10.06, p=0.002). Y-BOCS scores were higher in the OCD with depressive comorbidity group (mean 26.8 vs 24.2, t(119.66)=-4.12, p<.001), as were HAM-A scores (mean 18.6 vs 10, t(88.81)=-7.31, p<.001) and HAM-D scores (mean 14.2 vs 7.1, t(90.2)=-8.33, p<.001). Additionally, the prevalence of comorbid anxiety was higher in the OCD with depressive comorbidity group (60% vs 37.3%, $\chi 2(1)$=9.42, p=0.002)(Table S4).

The OCD with Anxiety group had an earlier age of OCD onset (mean 16.1 vs 18.1 years, t(237.74)=2.39, p=0.018), higher percentage of early-onset (63.7% vs 50.7%, $\chi 2(1)$=3.98, p=0.046), higher HAM-A scores (mean 14.3 vs 10.4, t(210.02)=-3.94, p<.001), higher HAM-D scores (mean 9.9 vs 8.0, t(203.85)=-2.44, p=0.015), and more prevalent comorbid depressive conditions (34.2% vs 17.1%, $\chi 2(1)$=9.42, p=0.002). Significant differences in DY-BOCS subdimensions included higher scores in Harm and Aggression (mean 6 vs 4.8, t(240.18)=-2.04, p=0.042), Symmetry and Ordering (mean 7.2 vs 4.7, t(239.42)=-4.77, p<.001), and Collecting and Hoarding (mean 1.8 vs 0.9, t(195.77)=-2.64, p=0.009) for the OCD with Anxiety group (Table S5).

#### 2.2. Obsessive-compulsive symptom dimensionality

##### *2.2.1. Cortical thickness and surface area*

The left rostral anterior cingulate thickness was positively associated with the sexual and religious dimension (B[SE]=0.01[0.004], p<0.001, $p_{(FDR)}$=0.061), and the right pericalcarine thickness had a negative association with the collecting and hoarding dimension (B[SE]=-0.02[0.01], p=0.004, $p_{(FDR)}$=0.261). The right rostral anterior cingulate and medial orbitofrontal areas showed positive associations with the sexual and religious dimension (right rostral anterior cingulate: B[SE]=0.01[0.004], p=0.020, $p_{(FDR)}$=0.691; right medial orbitofrontal: B[SE]=0.01[0.002], p=0.030, $p_{(FDR)}$=0.691). Additionally, an increase in the right parahippocampal thickness was associated with the collecting and hoarding dimension (B[SE]=0.02[0.01], p=0.048, $p_{(FDR)}$=0.968).

For cortical surface area, there was a negative association between the harm and aggression dimension and the right medial OFC (B[SE]=-13.11[3.68], p< .001), which remained significant after FDR correction ($p_{(FDR)}$=0.030). The sexual and religious dimension showed a positive association with the right entorhinal cortex (B[SE]=5.65[1.75], p=0.001), but this did not hold after FDR correction ($p_{(FDR)}$=0.098)(Table S20-S21).

*2.2.2. Subcortical*

The contamination dimension score was negatively associated with the accumbens area (B[SE]=-5.77[1.96], p=0.0036, $p_{(FDR)}$=0.042). Before multiple comparison corrections, scores on the harm and aggression dimension were negatively associated with pallidum volume (B[SE]=-7.87[3.68], p=0.0036, $p_{(FDR)}$=0.235) and cerebellar Crus I volume (B[SE]=-97.71[42.99], p=0.0239, $p_{(FDR)}$=0.163). For the sexual and religious dimension, positive associations were found with the volume of the amygdala (B[SE]=9.03[4.53], p=0.047, $p_{(FDR)}$=0.360), hippocampal head (B[SE]=10.64[3.86], p=0.0063, $p_{(FDR)}$=0.0165), hippocampal body (B[SE]=5.24[2.23], p=0.024, $p_{(FDR)}$=0.033), and whole hippocampal volumes (B[SE]=16.96[6.37], p=0.0083, $p_{(FDR)}$=0.0165)(Table 5 / Table S22-S26).

#### 2.3. Voxel-based Morphometry

*2.3.1. OCD vs HC*

In the VBM analysis comparing individuals with OCD > HC we observed no significant differences in GMV. In addition, no supra-threshold clusters were identified in the comparisons of HC > OCD, or in the SUIT analysis for both OCD > HC and HC > OCD contrasts (Table S146).

*2.3.2. OCD Severity*

Several clusters revealed significant negative associations, indicating decreased brain volumes with increased OCD severity in individuals with OCD. The primary findings included a significant cluster (t=4.32, Ke=1098, $P_{(FWE)}$=0.022, $MNI_{peak}$=8, 27, -21) spanning regions such as the right rectus, olfactory region, medial OFC, ventral striatum, and the left medial OFC. In the SUIT analysis a significant cluster was observed, demonstrating decreased cerebellar volumes associated with increased OCD severity.

This finding includes a cluster (t=3.99, Ke=156, $P_{(FWE)}$=0.004, $MNI_{peak}$=2, -56, -15) located in the cerebellar vermis 4 and 5 (Table S147/ Figure 2).

#### 2.3.3. *Obsessive-compulsive symptom dimensionality*

We did not observe any associations between GMV and OC symptom dimensions at the uncorrected cluster threshold of p<0.001, neither for VBM nor for SUIT analyses (Table S148).

#### 2.3.4. *Onset and duration of OCD illness*

We did not observe any differences in GMV between cases with early-onset and late-onset OCD for VBM and SUIT analyses (Table S149).

For duration of OCD illness, significant positive associations with brain volume were detected, revealing a cluster (t=5.5, Ke=1114, $P_{(FWE)}$=0.021, $MNI_{peak}$=18, 21, 50) involving the right superior frontal gyrus and right middle frontal gyrus. Additionally, a cluster (t=4.32, Ke=920, $P_{(FWE)}$=0.043, $MNI_{peak}$=21, -70, -10) was observed, implicating the right superior frontal gyrus (Table S150).

Conversely, in the SUIT cerebellar-specific analysis of duration of illness, significant negative associations were identified. A principal cluster (t=4.32, Ke=341, $P_{(FWE)}$<.001, $MNI_{peak}$=-30, -57, -60) was found in the left cerebellum lobule 8, extending to adjacent cerebellar areas. Another major cluster (t=4.29, Ke=1212, $P_{(FWE)}$<.001, $MNI_{peak}$=28, -67, -48) included the right cerebellum lobule 8, right cerebellum Crus 2, and right cerebellum 7b, indicating decreased cerebellar volumes with longer duration of illness. No supra-threshold clusters were found in the comparisons of late-onset > early-onset and vice versa, nor in the SUIT analysis for early-onset versus late-onset (Table S149-S150).

#### 2.3.5. *Comorbidity*

In the VBM analysis comparing individuals with OCD and with or without a comorbid depressive disorder, no significant clusters were observed. In the SUIT analysis, significant clusters were identified, revealing increased cerebellar volumes in the cases with comorbid depressive disorders. The primary finding includes a substantial cluster (t=4.56, Ke=516, $P_{(FWE)}$<0.001, $MNI_{peak}$=-37, -67, -37) covering areas of the left cerebellum crus 2, crus 1, lobule 8, and 7b. Another notable cluster (t=4.36, Ke=142, $P_{(FWE)}$=.006, $MNI_{peak}$=-33, -49, -23) was observed in the left cerebellum lobule 6 and the fusiform gyrus. Further findings reveal clusters at coordinates 5, -58, -9 (t=4.12, Ke=207, $P_{(FWE)}$=.001) involving the right cerebellum lobules 4 and 5, the Vermis, and the left cerebellum lobule 6; and at 2, -70, -37 (t=4.11, Ke=181, $P_{(FWE)}$=0.001) within the vermis 8 and left cerebellum lobule 8. Additional smaller clusters were identified at -9, -65, -50 (t=3.98, Ke=134, $P_{(FWE)}$=0.008) and -19, -58, -48 (t=3.89, Ke=106, $P_{(FWE)}$=0.024), predominantly in the left cerebellum lobule 8. A cluster at 4, -61, -40 (t=3.73, Ke=112, $P_{(FWE)}$=0.019) encompassed the vermis 9, vermis 8, and the left cerebellum lobule 8. When comparing cases with and without anxiety comorbidty, no differences were seen in GMV for VBM nor SUIT (Table S151).

##### *2.3.6. Past medication exposure*

We did not observe any differences in GMV between cases with and without prior SSRI/SNRI use, neither for VBM nor for SUIT analyses (Table S152).

#### ***2.4. Exploratory analyses***

##### *2.4.1. Cortical thickness and surface area with illness duration, anxiety and depressive symptom severity*

In the analysis of the association between cortical thickness and OCD illness duration, several regions demonstrated positive associations. Specifically, the right fusiform thickness showed a positive association (B[SE]=0.002[0.001], p=0.012, $p_{(FDR)}$=0.644), as did the left posterior cingulate (B[SE]=0.002[0.001], p=0.020, $p_{(FDR)}$=0.644), right rostral anterior cingulate (B[SE]=0.004[0.002], p=0.028, $p_{(FDR)}$=0.644), left insula (B[SE]=0.002[0.001], p=0.044, $p_{(FDR)}$=0.658), and right posterior cingulate thickness (B[SE]=0.002[0.001], p=0.047, $p_{(FDR)}$=0.658). However, these associations were not significant after FDR correction (Table S55/ Figure S6).

For surface area, the left transversetemporal cortex showed a positive association (B[SE]=1.39[0.52], p=0.008, $p_{(FDR)}$=0.511), indicating an increase in surface area with longer illness duration. The left frontal pole (B[SE]=0.63[0.27], p=0.019, $p_{(FDR)}$=0.581) and the right cuneus (B[SE]=4.18[1.86], p=0.026, $p_{(FDR)}$=0.581) also displayed a positive association. None of these associations remained significant after FDR correction (Table S56/ Figure S6).

HAM-D scores showed a negative association with cortical thickness in the right middle temporal cortex (B[SE]=-0.002[0.001], p=0.049), which was not significant after FDR correction ($p_{(FDR)}$=0.853). For surface area the right rostral anterior cingulate demonstrated a negative association (B[SE]=-2.88[1.00], p=0.004, $p_{(FDR)}$=0.301). A negative association was also observed in the right inferior temporal (B[SE]=-7.19[3.37], p=0.034, $p_{(FDR)}$=0.502) and right medial orbitofrontal regions (B[SE]=-3.13[1.52], p=0.041, $p_{(FDR)}$=0.502). However, after FDR correction, the associations did not remain significant (Table S57-58).

For HAM-A scores and cortical thickness no significant negative association was found. For surface area, the right rostral anterior cingulate cortex showed a significant negative association

(B[SE]=-2.73[0.76], $p<0.001$, $p_{(FDR)}=0.028$). Other regions such as the right inferior temporal (B[SE]=-5.60[2.57], $p=0.030$, $p_{(FDR)}=0.786$) and the right precuneus (B[SE]=-5.66[2.70], $p=0.037$, $p_{(FDR)}=0.786$) also showed negative associations, while the left transversetemporal cortex displayed a positive association (B[SE]=0.88[0.44], $p=0.046$, $p_{(FDR)}=0.786$). However, except for the right rostral anterior cingulate, these associations did not remain significant after FDR correction (Table S59-60).

#### *2.4.2. Subcortical and cerebellar volume*

No significant associations were found for subcortical volumes and duration of OCD illness. For hippocampal subfield segmentation a negative association was observed in the mean body of the hippocampus (B[SE]=-1.53[0.77], $p=0.048$), indicating a decrease in volume with longer OCD duration. However, this association was not significant after FDR correction ($p_{(FDR)}=0.192$). Cerebellar subfield segmentation analysis showed a positive association in mean lobule VIIIA (B[SE]=16.94[7.98], $p=0.035$), suggesting an increase in volume with longer OCD duration. However, these associations were not significant after FDR correction ($p_{(FDR)}=0.378$)(Table S61-65/ Figure S6).

For the association between HAM-D scores and subcortical volumes the right thalamus showed a significant association (B[SE]=11.58[5.47], $p=0.035$, $p_{(FDR)}=0.335$). Regarding subfield segmentations, we observed a significant association for the mean pulvinar volume (B[SE]=5.99[2.95], $p=0.044$, $p_{(FDR)}=0.108$)(Table S66-70).

In the analysis of HAM-A scores and their association with subcortical volumes, we observed a significant association with the right thalamus (B[SE]=10.36[4.17], $p=0.013$, $p_{(FDR)}=0.27$). For thalamic subfield segmentation we observed a positive associations with mean whole thalamus volume (B[SE]=17.99[7.20], $p=0.013$, $p_{(FDR)}=0.032$), as well as the ventral region (B[SE]=8.65[3.52], $p=0.015$, $p_{(FDR)}=0.032$), and the pulvinar (B[SE]=5.45[2.25], $p=0.016$, $p_{(FDR)}=0.032$)(Figure 3). No significant associations were observed in the cerebellum or other subcortical structures (Table 4/Table S71-75).

### 3. Supplementary TABLES

*Supplemental tables S6-S152 of the manuscript are available in the Zenodo repository. The tables provide comprehensive results from all freesurfer-based analyses, as well as voxel-based morphometry (VBM), conducted as part of the Global OCD consortium study.*

***Folder and Table Organization:***

- ***ComBat-harmonized FreeSurfer results*** *(Tables S6–S75):*
  [https://doi.org/10.5281/zenodo.15849703](https://doi.org/10.5281/zenodo.15849703)
  *This folder contains tables based on freesurfer analyses where site effects were harmonized using the ComBat approach.*

- ***Site-corrected FreeSurfer results*** *(Tables S76–S145):*
  [https://doi.org/10.5281/zenodo.15849783](https://doi.org/10.5281/zenodo.15849783)
  *This folder contains the same analyses, but with site included as a covariate rather than using ComBat harmonization.*

- ***Voxel-Based Morphometry (VBM) results*** *(Tables S146–S152):*
  [https://doi.org/10.5281/zenodo.15849703](https://doi.org/10.5281/zenodo.15849703)
  *This folder contains supplementary VBM cluster tables as described in the Results and Supplementary Methods.*

Table S1 – MRI scanners and parameters per site.

| *Site* | *Brazil* | *India* | *Netherlands* | *South Africa* | *U.S.A.* |
|---|---|---|---|---|---|
| *MRI Scanner* | Philips Achieva 3.0 T | Philips Ingenia 3.0 T CX | GE 3.0 T Discovery MR750 | Siemens MAGNETOM Skyra 3.0 T | GE 3.0 T SIGNA Premier |
| *Head coil* | 32-channel | 32-channel | 32-channel | 32-channel | 48-channel |
| *TR (ms)* | 6.5 | 6.5 | 6.9 | 2300 | 2235 |
| *TI (ms)* | 900 | 900 | 900 | 900 | 900 |
| *TE (ms)* | 2.9 | 2.9 | 3 | 2 | 2.8 |
| *Flip angle ($^{o}$)* | 9 | 9 | 9 | 9 | 9 |
| *Voxel size (mm)* | 1 x 1 x 1 | 1 x 1 x 1 | 1 x 1 x 1 | 1 x 1 x 1 | 1 x 1 x 1 |
| *Matrix* | 256 x 256 | 256 x 256 | 256 x 256 | 256 x 256 | 256 x 256 |

*Abbreviations: MRI, magnetic resonance imaging; TR, repetition time; TI, inversion time; TE, echo time.*

Table S2 – Demographic and clinical characteristics OCD onset

| Characteristic | Early-Onset (N=149) | Late-Onset (N=116) | *Statistics* |
|---|---|---|---|
| **Sex (N (%))** | | | χ2(1)=1.85, p=0.17 |
| Male | 75(50.3%) | 69(59.5%) | |
| Female | 74(49.7%) | 47(40.5%) | |
| **Age (years)** | 28.2 (7.8) | 31.2 (7.7) | t(248.95)=-3.12, p=0.002 |
| **Estimated IQ** | 106.8 (12.3) | 101.8 (12.3) | t(246.97)=3.26, p=0.001 |
| **Education (years)** | 15.2 (2.9) | 15.3 (2.6) | t(256.76)=-0.31, p=0.76 |
| **Y-BOCS** | 24.7 (4.5) | 25 (5.4) | t(224.08)=-0.46, p=0.65 |
| **Duration of illness**[a] | 15.8 (8.5) | 7.8 (6.2) | t(262.33)=8.8, p<0.001 |
| **Age of OCD onset (years)** | 12.5 (3.3) | 23.4 (5.6) | t(176.31)=-18.68, p<0.001 |
| **HAM-A** | 12.7 (8.4) | 11.4 (7.6) | t(257.59)=1.28, p=0.20 |
| **HAM-D** | 9 (5.9) | 8.6 (6.4) | t(237.06)=0.52, p=0.61 |
| **comorbid anxiety disorders** | 72(48.3%) | 41(35.3%) | χ2(1)=3.98, p=0.046 |
| **comorbid depressive disorders** | 40(26.8%) | 25(21.6%) | χ2(1)=0.72, p=0.40 |
| **Medication naïve (%)** | | | |
| SSRI | 84(56.4%) | 70(60.3%) | χ2(1)=0.27, p=0.60 |
| SNRI | 140(94%) | 112(96.6%) | χ2(1)=0.47, p=0.49 |
| Benzodiazepines | 133(89.3%) | 106(91.4%) | χ2(1)=0.13, p=0.71 |
| Antipsychotics | 132(88.6%) | 112(96.6%) | χ2(1)=4.63, p=0.031 |
| Mood stabilizers | 143(96%) | 115(99.1%) | χ2(1)=1.46, p=0.23 |
| CBT naïve | 104(69.8%) | 96(82.8%) | χ2(1)=5.24, p=0.022 |
| **DY-BOCS** | | | |
| Harm and Aggression | 5.3 (4.7) | 5.3 (4.7) | t(249.18)=-0.06, p=0.95 |
| Sexual and Religious | 4.5 (4.8) | 4.4 (5) | t(242.82)=0.19, p=0.85 |
| Symmetry and Ordering | 6.1 (4.4) | 5.2 (4.4) | t(247.47)=1.64, p=0.10 |
| Contamination | 6.4 (4.9) | 6.5 (5.2) | t(240.02)=-0.19, p=0.85 |
| Collecting and Hoarding | 1.4 (2.7) | 1.2 (2.6) | t(251.8)=0.68, p=0.50 |

Data are presented as mean (SD) unless otherwise indicated. a=duration of illness/age of OCD onset missing for 1 patient. Abbreviations: CBT=Cognitive Behavioral Therapy, Comorbid anxiety disorders=social anxiety disorder and/or generalized anxiety disorder, specific phobia, panic disorder, agoraphobia, posttraumatic stress disorder, comorbid depressive disorders=major depressive disorder or persistent depressive disorder, DY-BOCS: Dimensional Y-BOCS (severity score), HAM-A=Hamilton Anxiety Rating Scale, HAM-D=Hamilton Depression Rating Scale, SNRI=Selective Noradrenaline Reuptake Inhibitor, SSRI=Selective Serotonin Reuptake Inhibitor, Y-BOCS=Yale-Brown Obsessive-Compulsive Scale.

Table S3 – Demographic and clinical characteristics SSRI/SNRI naïve vs past SSRI/SNRI use

| Characteristic | **SSRI/SNRI naïve (N=151)** | **SSRI/SNRI history (N=115)** | *Statistics* |
|---|---|---|---|
| **Sex (N (%))** | | | χ2(1)=0, p=1 |
| Male | 69(45.7%) | 52(45.2%) | |
| Female | 82(54.3%) | 63(54.8%) | |
| **Age (years)** | 28.7 (7.8) | 30.6 (7.9) | t(244.07)=-1.92, p=0.056 |
| **Estimated IQ** | 105.4 (12.5) | 103.7 (12.5) | t(244.8)=1.12, p=0.26 |
| **Education (years)** | 15.2 (2.7) | 15.1 (2.9) | t(234.32)=0.28, p=0.78 |
| **Y-BOCS** | 24.2 (4.8) | 25.6 (5) | t(239.75)=-2.3, p=0.022 |
| **Duration of illness**[a] | 11.4 (8.7) | 13.5 (8.2) | t(252.1)=-1.97, p=0.05 |
| **Age of OCD onset (years)** | 17.3 (7.2) | 17.2 (6.8) | t(252.89)=0.2, p=0.84 |
| **OCD onset (N (%))**[a] | | | χ2(1)=0.92, p=0.34 |
| early-onset | 80(53.3%) | 69(60%) | |
| late-onset | 70(46.7%) | 46(40%) | |
| **HAM-A** | 11.1 (8) | 13.4 (7.9) | t(247.47)=-2.39, p=0.018 |
| **HAM-D** | 8 (5.6) | 9.8 (6.5) | t(227.05)=-2.41, p=0.017 |
| **comorbid anxiety disorders** | 57(37.7%) | 57(49.6%) | χ2(1)=3.26, p=0.071 |
| **comorbid depressive disorders** | 29(19.2%) | 36(31.3%) | χ2(1)=4.54, p=0.033 |
| **Medication naïve (%)** | | | |
| SSRI | 151(100%) | 4(3.5%) | χ2(1)=246.17, p<0.001 |
| SNRI | 151(100%) | 102(88.7%) | χ2(1)=15.6, p<0.001 |
| Benzodiazepines | 146(96.7%) | 94(81.7%) | χ2(1)=14.89, p<0.001 |
| Antipsychotics | 149(98.7%) | 96(83.5%) | χ2(1)=18.7, p<0.001 |
| Mood stabilizers | 149(98.7%) | 110(95.7%) | χ2(1)=1.3, p=0.25 |
| CBT naïve | 124(82.1%) | 77(67%) | χ2(1)=7.33, p=0.007 |
| **DY-BOCS** | | | |
| Harm and Aggression | 5.2 (4.5) | 5.5 (4.9) | t(231.45)=-0.55, p=0.58 |
| Sexual and Religious | 4.4 (4.7) | 4.5 (5.2) | t(226.64)=-0.23, p=0.82 |
| Symmetry and Ordering | 5.4 (4.4) | 6.2 (4.5) | t(239.23)=-1.42, p=0.16 |
| Contamination | 6.1 (4.8) | 6.9 (5.2) | t(230.89)=-1.36, p=0.18 |
| Collecting and Hoarding | 1.1 (2.5) | 1.5 (2.9) | t(217.27)=-1.1, p=0.27 |

Data are presented as mean (SD) unless otherwise indicated. a=duration of illness/age of OCD onset missing for 1 patient. Abbreviations: CBT=Cognitive Behavioral Therapy, Comorbid Anxiety=social anxiety disorder and/or generalized anxiety disorder, specific phobia, panic disorder, agoraphobia, posttraumatic stress disorder, comorbid depressive disorders=major depressive disorder or persistent depressive disorder, DY-BOCS: Dimensional Y-BOCS (severity score), HAM-A=Hamilton Anxiety Rating Scale, HAM-D=Hamilton Depression Rating Scale, SNRI=Selective Noradrenaline Reuptake Inhibitor, SSRI=Selective Serotonin Reuptake Inhibitor, Y-BOCS=Yale-Brown Obsessive-Compulsive Scale.

Table S4 – Demographic and clinical characteristics OCD with and without depressive comorbidity

| Characteristic | **OCD without depressive disorders (N=201)** | **OCD with depressive disorders (N=65)** | *Statistics* |
|---|---|---|---|
| **Sex (N (%))** | | | χ2(1)=10.06, p=0.002 |
| Male | 103(51.2%) | 18(27.7%) | |
| Female | 98(48.8%) | 47(72.3%) | |
| **Age (years)** | 29.6 (8) | 29.3 (7.8) | t(110.22)=0.27, p=0.79 |
| **Estimated IQ** | 104.8 (12.4) | 104.1 (13) | t(104.41)=0.38, p=0.70 |
| **Education (years)** | 15.2 (2.7) | 15.2 (2.9) | t(101.77)=-0.09, p=0.93 |
| **Y-BOCS** | 24.2 (4.9) | 26.8 (4.4) | t(119.66)=-4.12, p<0.001 |
| **Duration of illness**[a] | 12.3 (8.6) | 12.4 (8.5) | t(109.31)=-0.09, p=0.93 |
| **Age of OCD onset (years)** | 17.4 (6.7) | 16.9 (8) | t(95.07)=0.38, p=0.71 |
| **OCD onset (N (%))**[a] | | | χ2(1)=0.72, p=0.40 |
| early-onset | 109(54.5%) | 40(61.5%) | |
| late-onset | 91(45.5%) | 25(38.5%) | |
| **HAM-A** | 10 (6.6) | 18.6 (8.7) | t(88.81)=-7.31, p<0.001 |
| **HAM-D** | 7.1 (4.9) | 14.2 (6.3) | t(90.2)=-8.33, p<0.001 |
| **comorbid Anxiety** | 75(37.3%) | 39(60%) | χ2(1)=9.42, p=0.002 |
| **Medication naïve (%)** | | | |
| SSRI | 126(62.7%) | 29(44.6%) | χ2(1)=5.87, p=0.015 |
| SNRI | 193(96%) | 60(92.3%) | χ2(1)=0.77, p=0.38 |
| Benzodiazepines | 186(92.5%) | 54(83.1%) | χ2(1)=3.97, p=0.046 |
| Antipsychotics | 186(92.5%) | 59(90.8%) | χ2(1)=0.04, p=0.85 |
| Mood stabilizers | 197(98%) | 62(95.4%) | χ2(1)=0.5, p=0.48 |
| CBT naïve | 152(75.6%) | 49(75.4%) | χ2(1)=0, p=1 |
| **DY-BOCS** | | | |
| Harm and Aggression | 5 (4.5) | 6.2 (5.3) | t(95.84)=-1.66, p=0.10 |
| Sexual and Religious | 4.3 (4.9) | 4.9 (5) | t(107.52)=-0.85, p=0.40 |
| Symmetry and Ordering | 5.4 (4.3) | 6.8 (4.7) | t(100.87)=-2, p=0.049 |
| Contamination | 6.2 (4.9) | 7 (5.3) | t(101.37)=-1.07, p=0.29 |
| Collecting and Hoarding | 1 (2.3) | 2.3 (3.3) | t(85.5)=-3.03, p=0.003 |

Data are presented as mean (SD) unless otherwise indicated. a=duration of illness/age of OCD onset missing for 1 patient. Abbreviations: CBT=Cognitive Behavioral Therapy, comorbid anxiety disorders=social anxiety disorder and/or generalized anxiety disorder, specific phobia, panic disorder, agoraphobia, posttraumatic stress disorder, comorbid depressive disorders = major depressive disorder or persistent depressive disorder, DY-BOCS: Dimensional Y-BOCS (severity score), HAM-A=Hamilton Anxiety Rating Scale, HAM-D=Hamilton Depression Rating Scale, SNRI=Selective Noradrenaline Reuptake Inhibitor, SSRI=Selective Serotonin Reuptake Inhibitor, Y-BOCS=Yale-Brown Obsessive-Compulsive Scale.

Table S5 – Demographic and clinical characteristics OCD with and without Anxiety comorbidity

| Characteristic | **OCD without Anxiety disorders (N=152)** | **OCD with Anxiety disorders (N=114)** | *Statistics* |
|---|---|---|---|
| **Sex (N (%))** | 76(50%), 76(50%) | 69(60.5%), 45(39.5%) | χ2(1)=2.5, p=0.11 |
| Male | | | |
| Female | | | |
| **Age (years)** | 29.9 (7.9) | 29.1 (7.9) | t(244.59)=0.82, p=0.42 |
| **Estimated IQ** | 104.6 (12.7) | 104.7 (12.3) | t(247.22)=-0.08, p=0.94 |
| **Education (years)** | 15.1 (2.5) | 15.4 (3.1) | t(211.4)=-0.95, p=0.35 |
| **Y-BOCS** | 24.8 (4.9) | 24.8 (4.9) | t(244.38)=0.05, p=0.96 |
| **Duration of illness**[a] | 11.8 (8.9) | 13 (8) | t(253.4)=-1.22, p=0.22 |
| **Age of OCD onset (years)** | 18.1 (6.9) | 16.1 (7.1) | t(237.74)=2.39, p=0.018 |
| **OCD onset (N (%))**[a] | | | χ2(1)=3.98, p=0.046 |
| early-onset | 77(50.7%) | 72(63.7%) | |
| late-onset | 75(49.3%) | 41(36.3%) | |
| **HAM-A** | 10.4 (7) | 14.3 (8.8) | t(210.02)=-3.94, p<0.001 |
| **HAM-D** | 8 (5.2) | 9.9 (6.9) | t(203.85)=-2.44, p=0.015 |
| **comorbid depressive disorders** | 26(17.1%) | 39(34.2%) | χ2(1)=9.42, p=0.002 |
| **Medication naïve (%)** | | | |
| SSRI | 95(62.5%) | 60(52.6%) | χ2(1)=2.22, p=0.14 |
| SNRI | 147(96.7%) | 106(93%) | χ2(1)=1.23, p=0.27 |
| Benzodiazepines | 137(90.1%) | 103(90.4%) | χ2(1)=0, p=1 |
| Antipsychotics | 143(94.1%) | 102(89.5%) | χ2(1)=1.32, p=0.25 |
| Mood stabilizers | 149(98%) | 110(96.5%) | χ2(1)=0.15, p=0.70 |
| CBT naïve | 111(73%) | 90(78.9%) | χ2(1)=0.94, p=0.33 |
| **DY-BOCS** | | | |
| Harm and Aggression | 4.8 (4.6) | 6 (4.7) | t(240.18)=-2.04, p=0.042 |
| Sexual and Religious | 4.3 (4.9) | 4.6 (4.9) | t(240.91)=-0.56, p=0.58 |
| Symmetry and Ordering | 4.7 (4.2) | 7.2 (4.3) | t(239.42)=-4.77, p<0.001 |
| Contamination | 6.1 (5.1) | 6.8 (4.8) | t(250.12)=-1.21, p=0.23 |
| Collecting and Hoarding | 0.9 (2.3) | 1.8 (3) | t(195.77)=-2.64, p=0.009 |

Data are presented as mean (SD) unless otherwise indicated. a=duration of illness/age of OCD onset missing for 1 patient. Abbreviations: CBT=Cognitive Behavioral Therapy, comorbid anxiety disorders=social anxiety disorder and/or generalized anxiety disorder, specific phobia, panic disorder, agoraphobia, posttraumatic stress disorder, comorbid depressive disorders=major depressive disorder or persistent depressive disorder, DY-BOCS: Dimensional Y-BOCS (severity score), HAM-A=Hamilton Anxiety Rating Scale, HAM-D=Hamilton Depression Rating Scale, SNRI=Selective Noradrenaline Reuptake Inhibitor, SSRI=Selective Serotonin Reuptake Inhibitor, Y-BOCS=Yale-Brown Obsessive-Compulsive Scale.

### 4. Supplementary FIGURES

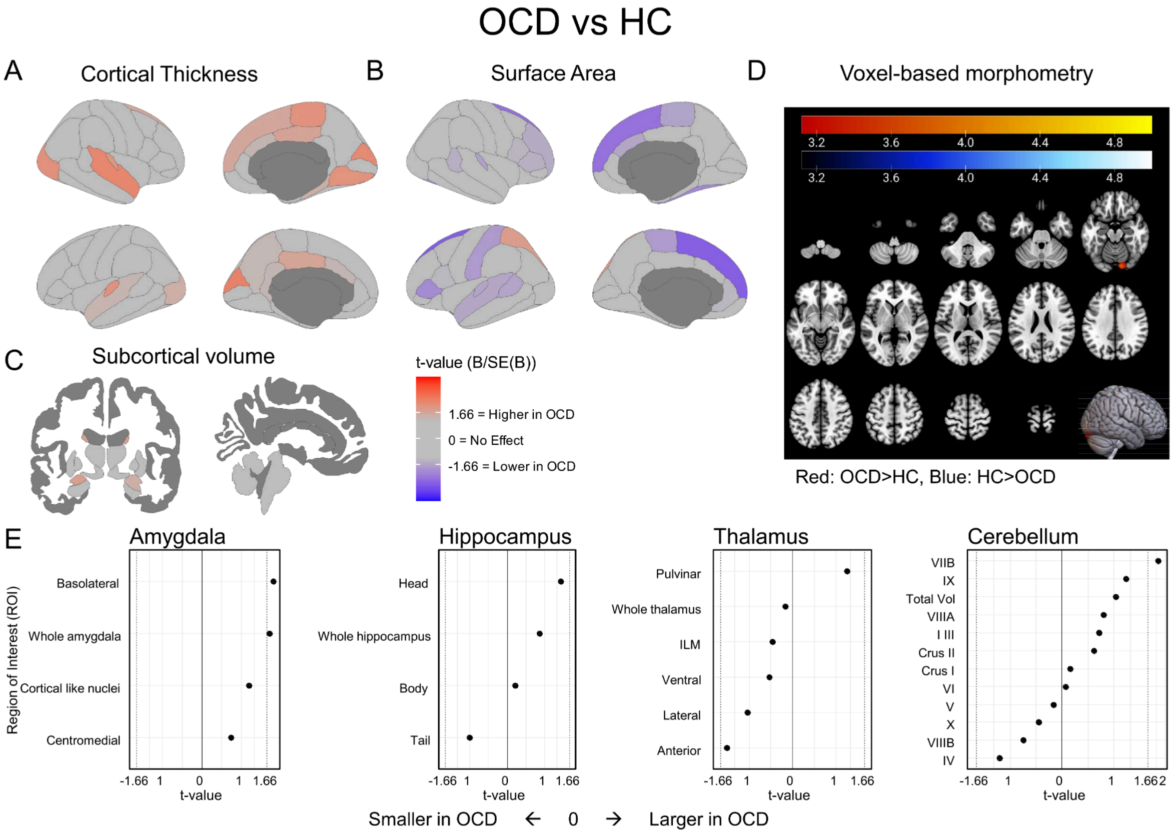


**Figure S1: ComBat-Corrected FreeSurfer and VBM findings for comparison between individuals with OCD (N=266) and Healthy Controls (N=254).** A: Cortical thickness maps with t-statistics from ComBat-corrected mixed models. B: Surface area maps with t-statistics from ComBat-corrected mixed models. C: Subcortical volume images with t-statistics from ComBat-corrected mixed models. D: Voxel-based morphometry results, shown at $p<0.001$ (uncorrected). E: Freesurfer subfield segmentations with t-statistics from ComBat-corrected mixed models; significance indicated by * ($p<0.05$, FDR-corrected).

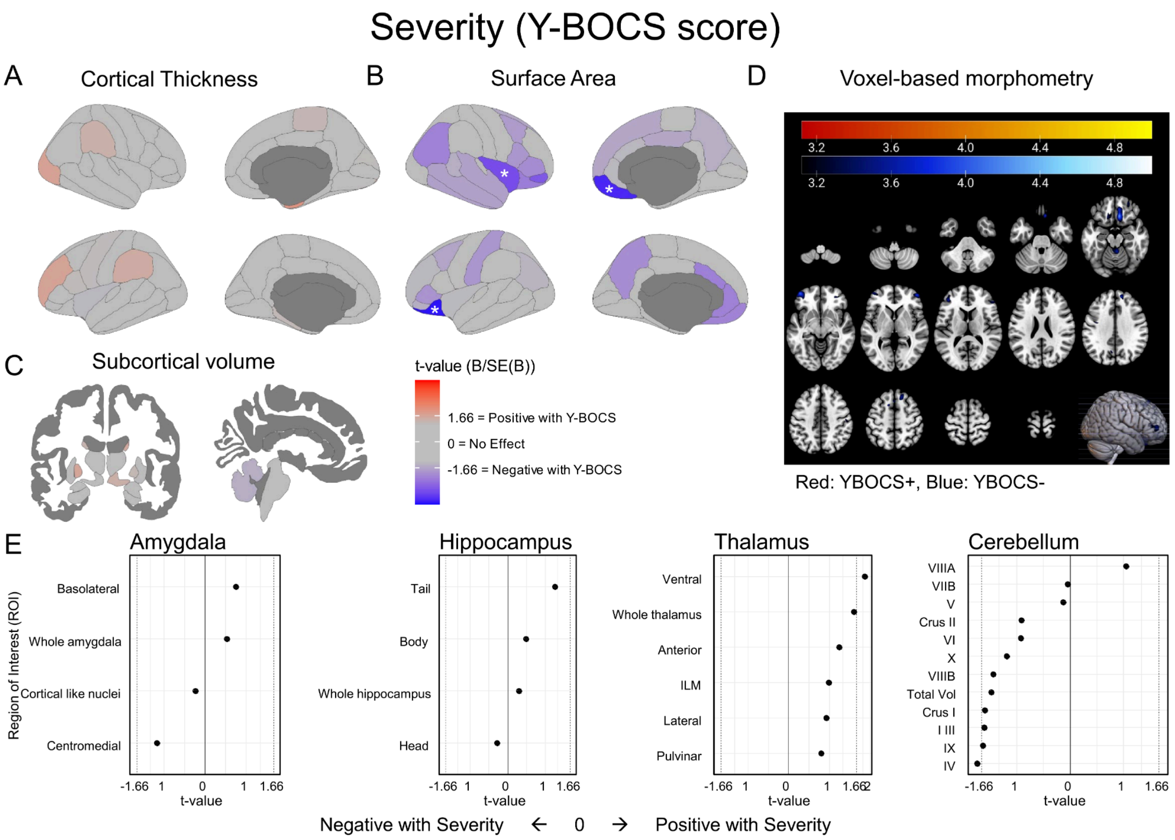


**Figure S2: ComBat-Corrected findings for association between FreeSurfer and VBM measures and OCD severity (N=266).** A: Cortical thickness maps with t-statistics from ComBat-corrected mixed models. B: Surface area maps with t-statistics from ComBat-corrected mixed models. C: Subcortical volume images with t-statistics from ComBat-corrected mixed models. D: Voxel-based morphometry results, shown at p<0.001 (uncorrected). E: Freesurfer subfield segmentations with t-statistics from ComBat-corrected mixed models; significance indicated by * (p<0.05, FDR-corrected).

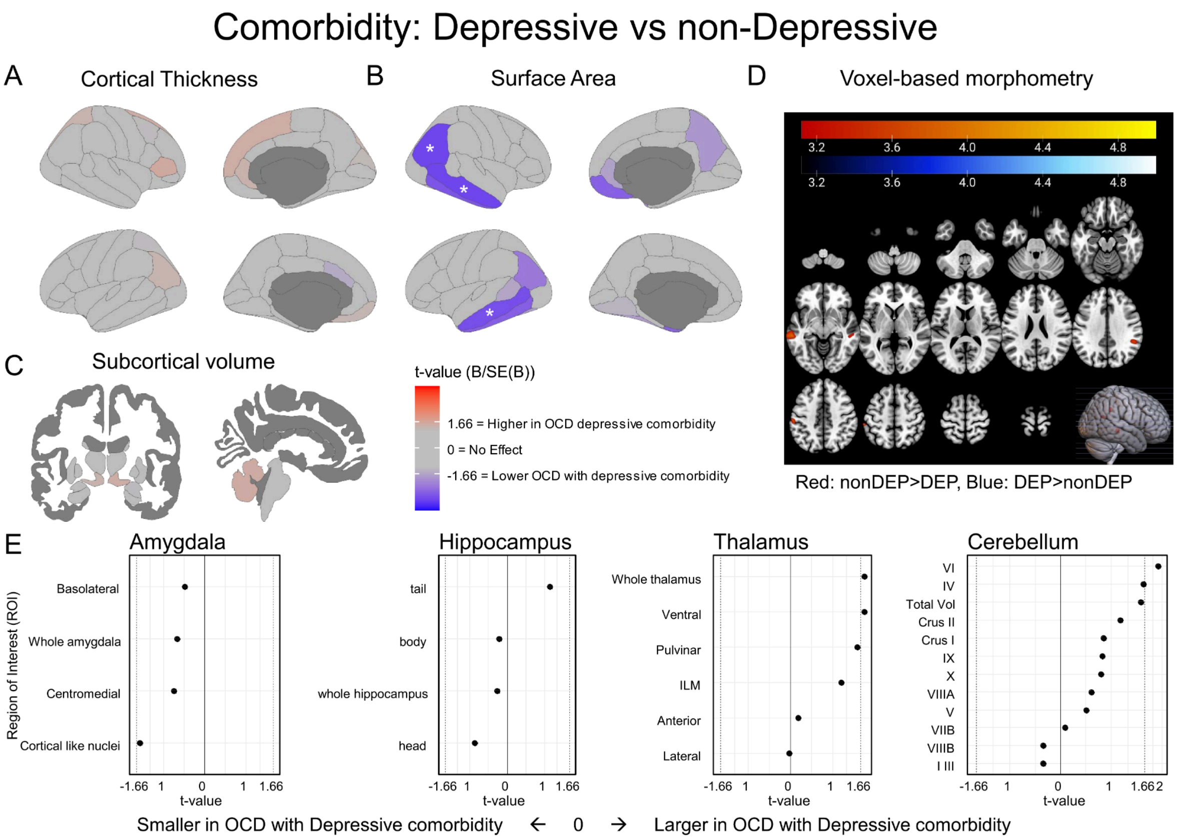


**Figure S3: ComBat-Corrected FreeSurfer and VBM findings for comparison between OCD cases with (N=65) vs without (N=201) depressive comorbidity.** A: Cortical thickness maps with t-statistics from ComBat-corrected mixed models. B: Surface area maps with t-statistics from ComBat-corrected mixed models. C: Subcortical volume images with t-statistics from ComBat-corrected mixed models. D: Voxel-based morphometry results, shown at p<0.001 (uncorrected). E: Freesurfer subfield segmentations with t-statistics from ComBat-corrected mixed models; significance indicated by * (p<0.05, FDR-corrected).

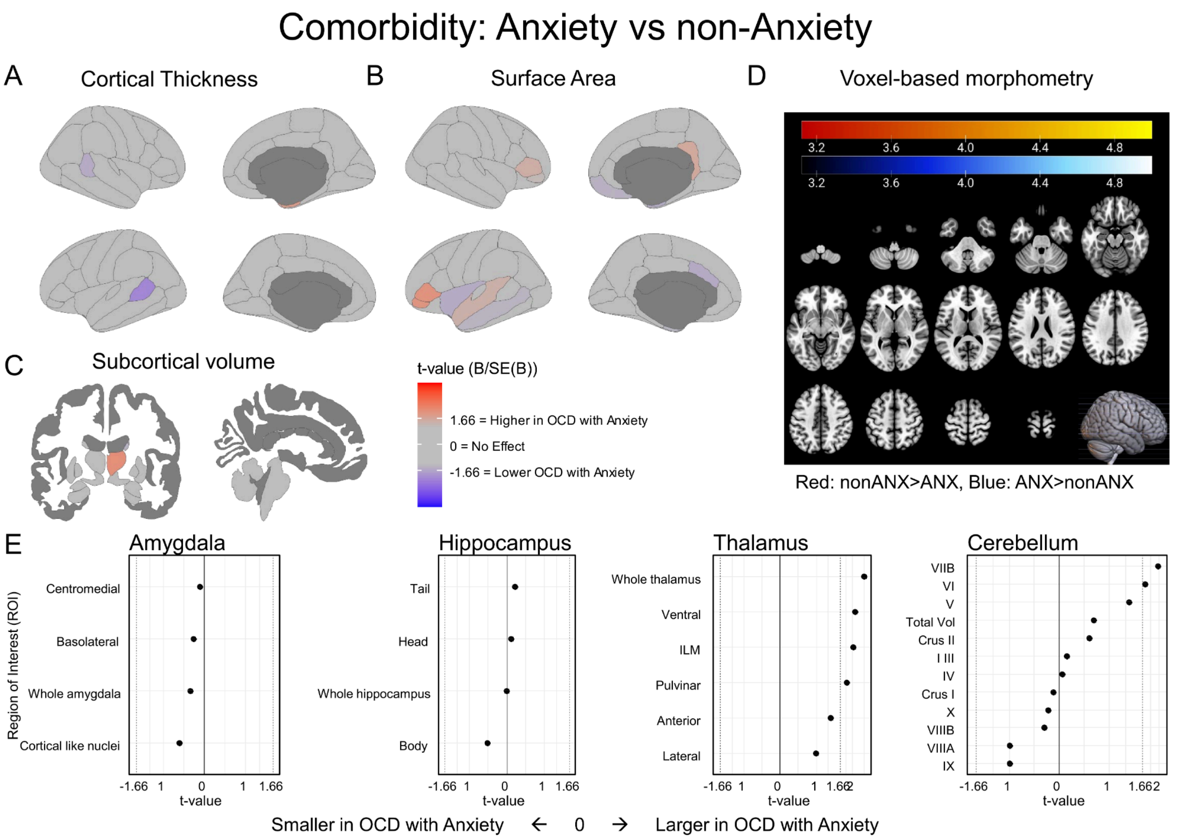


**Figure S4: ComBat-Corrected FreeSurfer and VBM findings for comparison between OCD cases with (N=114) anxiety vs without (N=152) anxiety comorbidity.** A: Cortical thickness maps with t-statistics from ComBat-corrected mixed models. B: Surface area maps with t-statistics from ComBat-corrected mixed models. C: Subcortical volume images with t-statistics from ComBat-corrected mixed models. D: Voxel-based morphometry results, shown at p<0.001 (uncorrected). E: Freesurfer subfield segmentations with t-statistics from ComBat-corrected mixed models; significance indicated by * (p<0.05, FDR-corrected).

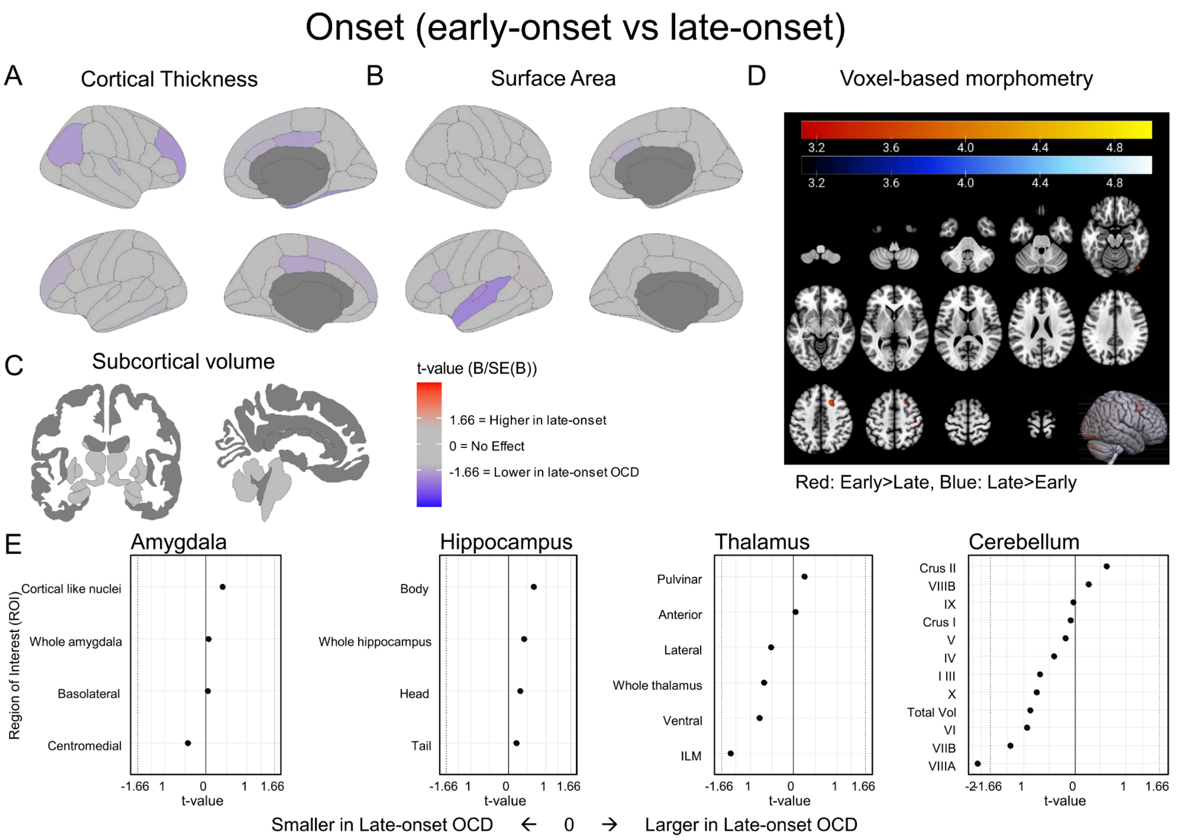


**Figure S5: ComBat-Corrected FreeSurfer and VBM findings for comparison between OCD cases with early (N=149) vs late (N=116) onset.** A: Cortical thickness maps with t-statistics from ComBat-corrected mixed models. B: Surface area maps with t-statistics from ComBat-corrected mixed models. C: Subcortical volume images with t-statistics from ComBat-corrected mixed models. D: Voxel-based morphometry results, shown at p<0.001 (uncorrected). E: Freesurfer subfield segmentations with t-statistics from ComBat-corrected mixed models; significance indicated by * (p<0.05, FDR-corrected).

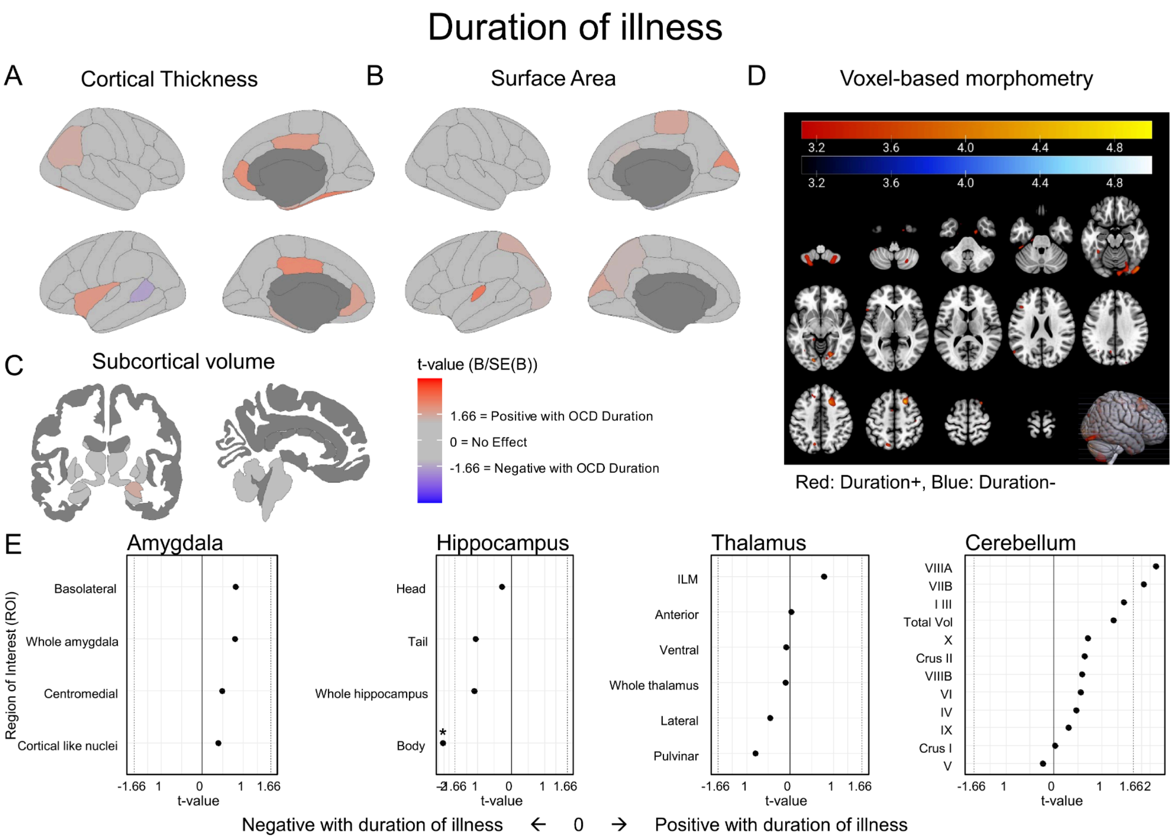


**Figure S6: ComBat-Corrected findings for association between FreeSurfer and VBM measures and duration of illness (N=266).** A: Cortical thickness maps with t-statistics from ComBat-corrected mixed models. B: Surface area maps with t-statistics from ComBat-corrected mixed models. C: Subcortical volume images with t-statistics from ComBat-corrected mixed models. D: Voxel-based morphometry results, shown at p<0.001 (uncorrected). E: Freesurfer subfield segmentations with t-statistics from ComBat-corrected mixed models; significance indicated by * (p<0.05, FDR-corrected).
